# Characterization of snRNA-related neurodevelopmental disorders through the Spanish Undiagnosed Rare Disease Programs

**DOI:** 10.1101/2025.09.16.25335449

**Authors:** Marta Sevilla-Porras, Esther Nieto-Molina, Zahara Medina, Alejandra Damián, Juan R. Tejedor, Carlos Ruiz-Arenas, Mario Cazalla, Rosario Carmona, Jorge Amigo, Ibai Goicoechea, Raúl Tonda, Gemma Bullich, Sergi Beltran, Noemí Toro-Barrios, Virginia Aquino, Emma Soengas-Gonda, Cinta Navarro-Moreno, Marcela Mena, June Corcuera, Verónica Martos-Gago, Míriam Álvarez-Barona, María Barreda-Sánchez, Cristina Silván, Jair Tenorio-Castaño, Carolina Alves, María I. Álvarez-Mora, Laia Rodríguez-Revenga, Irene Madrigal, María J. Ballesta-Martinez, Vanesa López-González, Belén Pérez, Montserrat Morales-Conejo, Irene Valenzuela, Marta Codina-Solà, Marta Alemany-Albert, Virginia Ballesteros-Cogollos, Raquel Rodríguez-López, Berta Almoguera, Isabel Lorda-Sánchez, Irene Lázaro-Rodríguez, Miguel A. Martín, Lidia Fernández-Caballero, Helena Gil-Peña, Noelia García-González, Marta Agúndez-Sarasola, Judith Armstrong, Loreto Martorell, Dídac Casas-Alba, Carmen Fons, Roser Urreizti, Antonio F. Martínez-Monseny, Leticia Pías-Peleteiro, Francisco Palau, Lexuri Gerrikabeitia, Nelmar V. Ortiz-Cabrera, Anna Duat-Rodríguez, Bárbara Fernández-Garoz, Laura López-Marín, Beatriz Bernardino-Cuesta, Elena Anton-Martin, María J. González-Gómez, Elena González-Alguacil, Carmen Gómez Lado, David Dacruz-Álvarez, Beatriz Sobrino, Víctor Martínez-Glez, Anna Ruiz, Carmen Manso-Bazús, Nino Spataro, Neus Baena, Juan Pablo Trujillo-Quintero, Nuria Capdevila, Anna Brunet-Vega, Eugenio Zapata-Aldana, Verónica A. Seidel, Raquel Yahyaoui, Ignacio Blanco, Elisabeth Castellanos, Montse Pauta, María del Mar Rovira, Barbara Masotto, Agustí Rodríguez-Palmero, Aurora Pujol, Agatha Schlüter, María Palomares-Bralo, María Á. Gómez-Cano, María Nieves-Moreno, Emi Rikeros-Orozco, Antonio Poyatos-Andujar, Inmaculada Medina-Martínez, Fernando Santos-Simarro, Damià Heine-Suñer, Susana R. Avella-Klaassen, Ángeles Perez-Granero, María Antonia Grimalt, Ignacio Arroyo-Carrera, Andrea Sariego-Jamardo, Ana I. Vega, José L. Fernández-Luna, Mireia Del Toro, Flora Sánchez-Jiménez, Juan M. Borreguero-León, Andrea Campo-Barasoain, Joaquín Dopazo, Mario F. Fraga, Feliciano Ramos, Jordi Rosell, Enrique Galán-Gomez, Salud Borrego, Luis Castaño, Francisco Barros, José M. Millán, Gemma Aznar-Laín, Encarna Guillén-Navarro, Carmen Ayuso, Ángel Carracedo, Pablo Lapunzina, Beatriz Morte, Luis A. Pérez-Jurado

## Abstract

Programs for Undiagnosed Rare Diseases (URD) with anonymized data sharing are contributing to the earlier genetic diagnosis of patients and to the identification and characterization of novel genetic disorders. Recently, *de novo* pathogenic variants in two non-coding spliceosomal small nuclear RNAs (snRNAs), *RNU4-2* and *RNU2-2*, key regulators of gene expression during neurodevelopment, have been linked to neurodevelopmental disorders (NDD). With the aim to identify individuals with causal genetic variants in known and novel snRNA genes, we analyzed WGS and clinical data of 1,708 probands with undiagnosed NDD enrolled in the Spanish URD Programs. Selected individuals underwent in-depth re-phenotyping including facial mask analyses (Face2Gene), and blood-DNA methylation profiling was obtained (Infinium methylation Epicv2 microarrays). We identified 38 patients (2.22%) with *de novo* pathogenic variants in snRNA genes, 28 in *RNU4-2* (1.64%), 6 in *RNU2-2* (0.35%), and 4 in *RNU5B-1* (0,23%). A clinically recognizable NDD (ReNU syndrome) is caused by *RNU4-2* variants, with dysmorphic features making a disease-associated facial mask, *RNU2-2* associates an early onset epileptic encephalopathy, and *RNU5-B1* a less defined NDD with overlapping features. A DNA methylomic episignature was also identified for *RNU4-2*, which does not overlap with the other conditions. We have also defined autosomal recessive inheritance in 8 additional patients (0.47%) with NDD and biallelic variants in three snRNA genes (3 in *RNU4-2*, 3 in *RNU2-2*, and 2 in *RNU4-1*). Our study has facilitated the quick diagnosis, management, and genetic counselling of 2.69% previously undiagnosed patients with NDD, further reinforcing the value and utility of the URD Programs.

**GRAPHICAL ABSTRACT:** 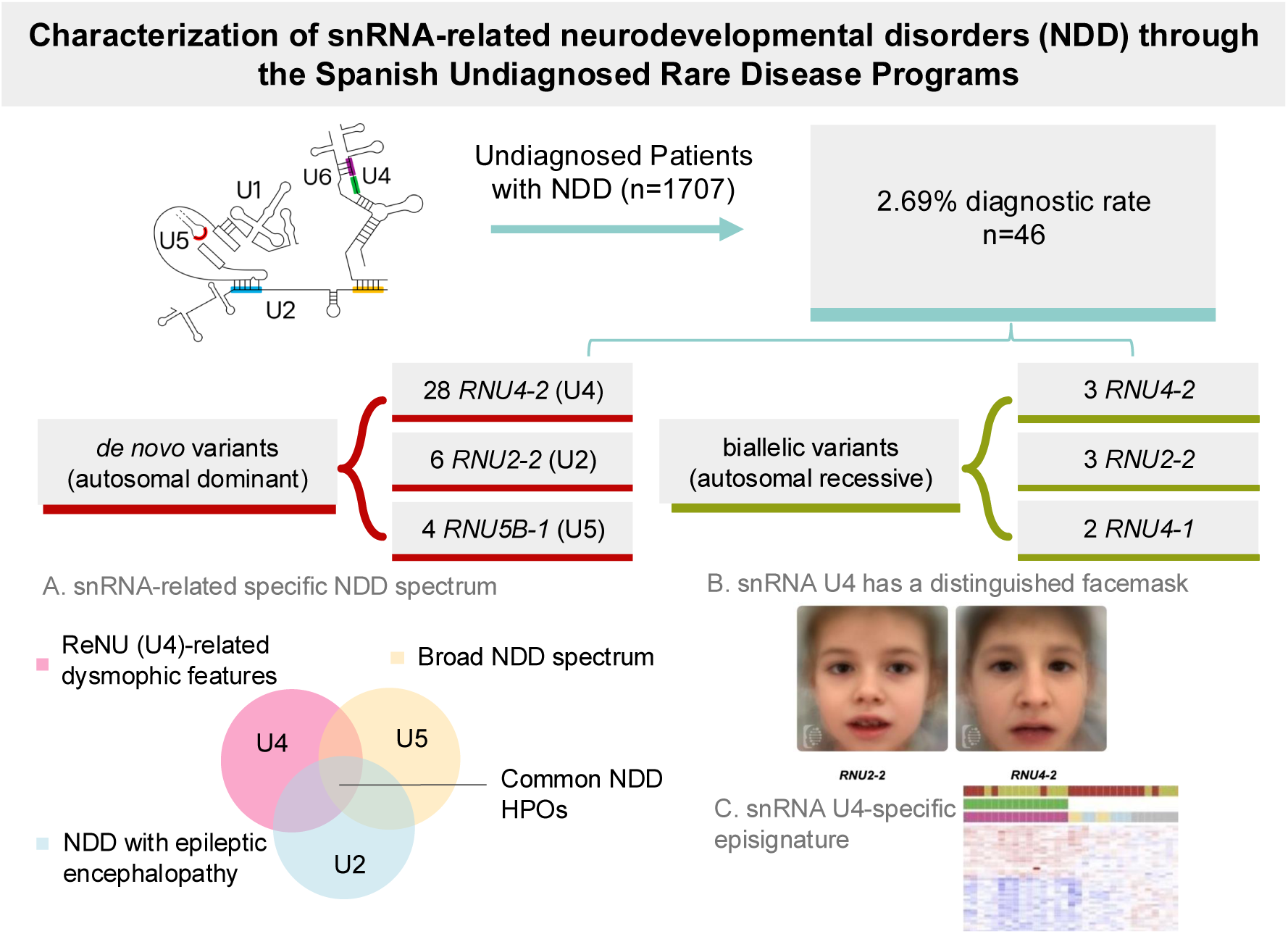

## INTRODUCTION

Patients with undiagnosed rare diseases, often of genetic origin, impose a remarkable health burden that stands at the intersection of research and clinical practice. Recognizing this gap, the International Rare Diseases Research Consortium (IRDiRC) has prioritized accelerating diagnostic solutions through the implementation of genomic medicine. In Spain, several national and regional programs coordinated through the CIBERER consortium^1^ have been launched since 2015: ENoD-CIBERER (https://www.ciberer.es/plataformas-ciberer/enod), URD-Cat (https://www.cnag.eu/projects/urdcat), and IMPaCT-Genómica (https://genomica-impact.es/). Aimed at advancing the diagnostic process beyond standard clinical care and guided by principles of equity, a network of 100 hospitals has been established across the country. These initiatives integrate standardized phenotypic and genomic data from large cohorts of undiagnosed patients to support rapid collaborative reanalysis and facilitate the discovery of novel disease-associated genes^2^.

A step forward in the diagnostic process has been made with whole genome sequencing (WGS), which enables the identification of multiple types of genetic variants and complex structural rearrangements and expands the scope to non-coding regions, including regulatory elements and non-coding genes. In the last year, emerging studies have highlighted small nuclear RNA (snRNA) genes, core components of the major spliceosome also referred as U-RNAs, as highly prevalent causes of neurodevelopmental disorders (NDDs) accounting for approximately 2% of cases. Given the genetic complexity of NDDs and the high rate of undiagnosed individuals, this represents a major advance. Other than fragile X syndrome, no single genes are responsible for more than 0.5% of NDD cases^3^.

snRNAs function within the spliceosome, a ribonucleoprotein complex essential for pre-mRNA splicing, and disruptions in their sequence or structure can impair normal splicing and gene expression. At least four snRNA components of the major spliceosome, *RNU2-2* (OMIM#621238), *RNU4-2* (OMIM#620823), *RNU5A-1* (OMIM#180691) and *RNU5B-1* (OMIM#621090), have recently been implicated in NDDs^4–8^. In addition, pathogenic variants in spliceosomal proteins such as *U2AF2, RBFOX1* and *PRPF19* which interact with these snRNAs within the U2/U4/U5 tri-SRNP complex, have also been reported in individuals with neurodevelopmental disorders^9^. The discoveries of snRNA genes were driven by the fact that pathogenic variants in those genes tend to cluster within functionally conserved regions that show notable depletion of variation in the population, supporting their evolutionary constraint and pathogenic potential. In *RNU4-2*, pathogenic variants cluster within an 18-bp region critical for U4-U6 snRNA interaction and spliceosome function, causing a distinct and recurrent neurodevelopmental syndrome termed ReNU (OMIM#620851)^7,10,11^. Similarly, pathogenic variants in *RNU2-2*, a gene initially annotated as a pseudogene but now recognized as a functional and highly expressed gene^12,13^, alter critical motifs involved in branch point recognition and spliceosomal assembly^5^. Further studies have expanded the mutational landscape to include *RNU5B-1* and *RNU5A-1*, where variants cluster in the conserved loop I domain, a region responsible for aligning the 5′ and 3′ exon ends during the splicing reaction^7,8,14,15^. Functional studies support the pathogenicity of these snRNA mutations. In particular, transcriptomic analyses on blood samples or fibroblasts from patients with *RNU4-2* variants show quantitatively altered splice-site usage in a significant proportion of transcripts^6,7^. However, unlike disorders linked to snRNA components of the minor spliceosome, such as *RNU4ATAC* or *RNU12*, widespread intron retention has not been consistently observed in affected individuals, suggesting a distinct splicing defect mechanism^5,11^.

In this study, we report the identification of *de novo* pathogenic variants in the constraint regions of three snRNA genes (*RNU2-2*, *RNU4-2* and *RNU5B-1*) in 38 of 1,708 probands with undiagnosed NDDs enrolled in Spanish URD programs. We have also defined biallelic inheritance of rare variants outside the constraint regions of three snRNA genes (*RNU2-2*, *RNU4-1* and *RNU4-2*) in 8 additional individuals with NDD including two siblings, supporting the growing evidence for autosomal recessive inheritance of RNU-associated disorders^1617^. Through comprehensive phenotypic characterization, we delineate gene-specific clinical profiles, including a distinctive facial gestalt in *RNU4-2*-related (ReNU) syndrome, identified using facial recognition tools (Face2Gene), and a domain-specific DNA methylation episignature uniquely associated with *RNU4-2* variants. We have also explored the possibility that oligogenic models involving different snRNA subunits may underlie disease in a subset of cases, searching for combinations of damaging variants present in patients more often than would be expected by chance. Overall, we have achieved a final diagnosis for 2.69% of the enrolled patients with previously undiagnosed NDD, with immediate implications for management and genetic counseling. Our findings reinforce the relevance of snRNA mutations and spliceosome regulation in the etiology of NDDs and underscore the clinical value of national URD programs.

## SUBJECTS AND METHODS

### Patients enrolled in Spanish URD Programs

Inclusion criteria for patients in the Spanish URD Programs (IMPaCT-Genómica, URD-Cat and ENoD-CIBERER) included: 1) a suspected genetic disorder that remained undiagnosed; 2) a complete diagnostic assessment with exome sequencing, now considered the gold standard among genetic testing; 3) reanalysis of exome data if analysis was done >1 year ago (most cases); and 4) informed consent to the storage and anonymous sharing of clinical and genomic data. There are no age or disease-type restrictions, but most recruited cases are congenital or pediatric-onset, and predominantly with neurological disorders.

The aggregate data of these programs (until 12/2024) comprised 2,200 probands and 1,000 unaffected relatives (mostly parents), including 1,708 probands with NDDs. Phenotypic information from all patients has been collected on a platform developed specifically for these programs, with standardized Human Phenotype Ontology (HPO)^18^ terms to ensure consistent clinical data collection and analysis. An average of 11.3 terms per case was recorded.

### Whole genome sequencing and data processing

Methods for whole genome sequencing in the URDCat and ENoD projects has been previously reported (https://www.ciberer.es/plataformas-ciberer/enod)^19^. For cases recruited in IMPaCT-Genómica, 1 µg of DNA was used for library preparation using the KAPA HyperPrep/ HyperPlus Kit (Roche Sequencing Solutions, Inc.) following manufactureŕs protocol. WGS libraries were sequenced on the NovaSeq 6000 or NovaSeq X Plus (Illumina, Inc.) with 2×150 bp pair-end reads, achieving ≥30X mean coverage, >400 bp mean insert size, and >15X depth for ≥95% of the GRCh38. Generated raw sequencing data was processed either through Illumina DRAGEN Bio-IT Platform (v4.0-v4.3) or through benchmarked standard analysis pipelines that used BWA-MEM or BWA-MEM2^20,21^ for read alignment to version 38 of the human genome and GATK HaplotypeCaller (v4.1-4.3) for the detection of single nucleotide variants (SNV) and short insertions and deletions in line with the GATK Best Practices workflow for germline short variant Discovery^22^. Structural variants were detected using Manta, Control-FREEC or GRIDSS^23–25^ and expansions of short tandem repeats (STRs) were identified via ExpansionHunter^26^. All runs of homozygosity larger than 500kb in length were identified, and read depth calculated across the target region of interest to provide a measure of data quality. In addition to comprehensive individual case analysis and reanalysis, genomic data of the entire dataset and/or subsets with similar phenotypes can also be analyzed to identify all individuals with variants in specific genes or regions. The annotated genomic variants of URD-Cat and part of EnOD and IMPaCT was available in databases or platforms such as GPAP^26^. These databases enable to query for specific variants, regions or genes through a Graphical User Interface or an Application Programming Interface. In other cases, genomic variants were available in VCF and other text format files.

### Ethics statement

Participants were enrolled through the clinical services of participating Spanish hospitals under protocols subsequently approved by the Instituto de Salud Carlos III Research Ethics Committee (CEI-PI01_2022) and endorsed by the institutional review boards of the participating centers. Written informed consent was obtained from all participants at pre-test counseling sessions, allowing the sharing of pseudonymized clinical information with international collaborators and researchers. Additional written consent to publish clinical data and photographs was also collected in accordance with local ethical guidelines and best practices.

### Analysis of candidate variants in snRNA genes

We screened the three cohorts of undiagnosed patients in the Spanish URD Programs for potentially pathogenic variants in the *RNU4-2* (NR_003137.2) and *RNU2-2* (NR_199791.1) genes, previously associated with NDDs. RD-Cat GPAP and CNAG GPAP, both based on RD-Connect GPAP^19^, were used to query the datasets, specifically leveraging the “search across all” functionality to efficiently identify candidate variants. We also analyzed structural variants and small insertions or deletions (indels) within a ±2000 base-pair window flanking either of the two loci. Given that the previously reported pathogenic variants in these genes were found to cluster in regions depleted of variation in the population, we extended our analysis to other small nuclear RNA (snRNA) genes of the major spliceosome that are similarly constrained. A threshold of −0.25 in the normalized observed proportion of single nucleotide variants (SNVs) was used to define such regions of constraint^6^. Only two additional snRNA genes, *RNU5B-1* (NR_002757.3) and *RNU4-1* (NR_003925.3), contained regions meeting this criterion. Variants were annotated with allele counts (AC) from gnomAD v4.1.0, retaining those with AC < 40 for further analysis. All candidate variants were visually reviewed using the Integrative Genome Viewer (IGV)^27^. Selected variants were categorized by type, population frequency, and inheritance pattern.

We also investigated the potential contribution of biallelic variants in snRNA genes to NDD, assessing whether such biallelic variants are enriched in affected individuals (probands) with respect to unaffected relatives. This analysis also focused on snRNA genes constituting the major spliceosome subunits (U1, U2, U4, U5, U6), curated based on their expression profiles in brain tissues using Genotype-Tissue Expression (GTEx) data. Variants were filtered based on population frequency, retaining those with an allele frequency (AF) <0.001 in gnomAD v4.1.0 and an internal cohort frequency <0.002. Manual curation was performed using IGV to evaluate read-level evidence and determine the phasing of compound heterozygous variants (*cis* versus *trans* configuration). We selected biallelic configurations, defined as either (i) homozygous or (ii) compound heterozygous variants confirmed to be in *trans*. Additionally, we evaluated the enrichment of digenic combinations, comparing the frequency of individuals carrying two or more rare variants in different snRNA genes. For these digenic analyses, we applied the same filtering and selection criteria as described for biallelic variants. Comparisons were then performed between the NDD proband cohort (n=1708) and controls (n=2121, unaffected relatives and non-NDD cases) to assess whether digenic configurations were enriched in cases.

Sanger sequencing following PCR amplification of the target regions was performed in trios to confirm inheritance. PCR primers used to amplify genomic DNA are detailed in **Table S1**.

### RNA folding prediction

NR_199791.1 (*RNU2-2*), NR_003925.2 (*RNU4-1*), NR_003137.3 (*RNU4-2*) and NR_002757.3 (*RNU5B-1*) sequences were used as templates to predict the RNA secondary structure of the respective snRNAs via RNAfold^28^. Predictions were generated for both the wild-type RNA and mutant versions carrying selected putative pathogenic variants identified in our cohort. Default parameters were applied: Turner model 2024, avoidance of isolated base pairs, and dangling energies applied on both sides of a helix. A folding constraint was applied only for *RNU5B-1* to preserve known structural features, specifically preventing base pairing at the nine nucleotides of the Sm-binding site^29^.

### Reverse phenotyping

Clinical reevaluation was conducted at each participating institution for all patients carrying *de novo* pathogenic variants in *RNU2-2*, *RNU4-2* or *RNU5B-1*, to ensure accurate and standardized phenotypic data collection. A total of 200 individual HPO terms were collected from the phenotypic data of the 38 patients, and clinicians (neurologist, neuropediatrician and/or clinical geneticist) were responsible for validating and expanding phenotypic annotations. To evaluate phenotypic variability among patients with variants in the three gene groups, we performed unsupervised hierarchical clustering using HPO-coded features. Patients with incomplete phenotypic annotation (<19% features) were excluded, resulting in a final cohort of 32 individuals: 22 with *RNU4-2*, 6 with *RNU2-2*, and 4 with *RNU5B-1* variants. Clustering was based on Euclidean distance and Ward’s linkage, and results were visualized through a heatmap to illustrate shared and distinct phenotypic features.

We compared the frequency of individual HPO terms across genes to identify gene-specific phenotypic signatures. HPO terms were considered gene-specific if they showed ≥30% difference in frequency between genes and occurred in >50% of patients for the gene of interest.

### Face-mask analysis

Frontal facial photographs of individuals with *de novo* dominant mutations in *RNU4-2*, *RNU2-2* and *RNU5B-1* genes were analyzed using the Face2Gene Research application tool (FDNA Inc., USA). The analysis included 15 individuals with *RNU4-2* variants, 10 with *RNU2-2* variants, 4 with *RNU5B-1* variants, and a comparison cohort of 31 unaffected individuals matched for sex, ethnicity, and age distribution (**Table S2**). Two main algorithms were used in these analyses: DeepGestalt and GestaltMatcher ^30,31^. The DeepGestalt algorithm pre-processes facial images by detecting landmarks, aligning the face, and analyzing specific regions through a Deep Convolutional Neural Network (DCNN). It generates a softmax vector indicating correspondence to known syndromes and creates a ranked list of possible syndrome matches, providing a DeepGestalt (DG) score for each suggested syndrome. Additionally, DeepGestalt enables binary comparisons between cohorts, calculating Area Under the Curve (AUC) values for the ROC curve, to quantify group distinguishability. The second algorithm applied is GestaltMatcher, which encodes patient photos into 320-dimensional Facial Phenotype Descriptors (FPDs) and compares the resulting vectors to each other creating a “Clinical Face Phenotype Space”, such that the distances between photos define syndromic similarity. GestaltMatcher can determine similarity between query images or to individually embedded images, provide syndrome suggestions for single query images, and visualize clustering patterns using t-distributed Stochastic Neighbor Embedding (t-SNE) plots.

### Genome-wide DNA methylation profiling

DNA samples were extracted from whole blood of 16 affected individuals with *RNU4-2* variants, 5 individuals with *RNU2-2* variants and 4 individuals with *RNU5B-1* variants using standard protocols. Patients’ samples, along with DNA samples from 47 age- and sex-matched controls were processed and hybridized to the Illumina Infinium MethylationEPICv2 BeadChip arrays (San Diego, CA, USA) following the manufacturer’s protocol. The resulting intensity data files were processed in R (version 4.2.3) using the meffil package^32^. Additional control data were obtained from GSE accession number GSE246337 (45 samples) (**Table S3**). Stringent quality control (QC) measures were applied to ensure data integrity. Samples and CpG sites were filtered based on bead count and detection *p-value* thresholds (both set at 0.05). To ensure data integrity, we used the xreactive_probes function from the minfi package^33^ to identify and exclude probes according to established stringent quality-control criteria. In addition probes that met some of this criteria were excluded: 1) probes located on sex chromosomes (chrX or chrY), 2) probes containing common SNPs at the CpG site (population frequency >1%), 3) probes located at single nucleotide extension sites (masked probe list from https://github.com/zhou-lab/InfiniumAnnotationV1), 4) probes non-CpG targeting and 5) probes known to cross-hybridize with off-target chromosomal sites^33^. Sex prediction was assessed with meffil package using a standard deviation threshold of 3. Normalization was carried out using functional normalization^34^. A selection of 3-5 principal components was determined based on residual analysis to optimize data normalization. Every pool of samples was normalized separately. Finally, to address technical variations, batch effect correction was performed using the ComBat function from the sva package^35^, effectively mitigating array-related batch effects.

### Mapping of DNA methylation episignatures

For *RNU4-2*, a subset of 10 affected individuals was selected as the training cohort to map DNA methylation signatures and train classification models. The remaining 6 individuals were used as a testing dataset to evaluate the classification model’s performance (**Table S3**). A matched control group, four controls per case, was selected using the MatchIt package for the training cohort^36^. Matching was based on predicted age (calculated with the DNAmAge function from the methylclock package)^37^ and sex^32^. For *RNU2-2* and *RNU5B-1*, due to the small sample size, no formal distinction between training and validation groups was made, and all cases were analyzed together.

Beta values, ranging from zero (no methylation) to one (full methylation), were transformed into M-values using a logit transformation. Linear regression analysis was performed using the limma package^38^ to detect differentially methylated probes (DMPs). Estimated blood cell proportions (blood gse167998^32^), age and sex were included in the model matrix to account for confounding variables. P-values were moderated using the eBayes function and corrected with the Benjamini and Hochberg (BH) algorithm.

We employed a three-step probe selection strategy, adapted from a previous publication^39^:

1. Probes were ranked based on the product of mean methylation difference (Δβ) and the negative logarithm of the corrected p-value, using the formula −|Δβ| log p, with a pre-selection that retained only probes passing an adjusted p<0.05 and displaying an absolute Δβ≥0.05, thus ensuring both statistical significance and a biologically meaningful effect size.
2. AUROC values were calculated for the top 1000 probes, and the 500 probes with the highest classification performance were retained.
3. Highly correlated probes with pairwise Pearson correlation coefficients >0.75 were removed to avoid redundancy.

The final selection of probes was assessed using hierarchical clustering and multidimensional scaling (MDS) to confirm robust differentiation between cases and controls. The reproducibility of the episignature was further validated using leave-one-out cross-validation (LOOCV). A support vector machine (SVM) classifier was trained using the final probe set. The model was trained using a radial basis function (RBF) kernel. The SVM implementation was performed using the e1071 package^40^ with a fixed random seed to ensure reproducibility.

### Detection of Differentially Methylated Regions (DMRs)

Differentially methylated regions (DMRs) were identified using the DMRcate package^41^. Regions containing at least 5 significantly different CpGs within 2,5 kb, with a minimum mean methylation difference of 10% and a Fisher’s multiple comparison *p-value* <1e-4, were considered significant. To further characterize these DMRs, we used UCSC Genome Browser tracks: Refseq Genes, CpG Islands, the H3K27Ac Mark from the ENCODE regulation track, and ENCODE GeneHancer regulatory elements.

### Functional enrichment analysis of DMPs

For enrichment analysis, we used the top 500 probes with the highest AUROC performance, as identified in the feature selection pipeline. This approach ensured that the probes included in the functional analysis were those with the strongest discriminatory power between cases and controls, maximizing the detection of biologically relevant pathways. To assess the biological relevance of DMPs and near-by genes, we conducted gene-set enrichment analysis (GSEA) using the missMethyl package^42^. The full set of analyzed CpGs (647,153 probes) was used as the background to ensure robust statistical inference. Gene Ontology (GO) enrichment was performed using the gometh function, analyzing Biological Process (BP), Cellular Component (CC), and Molecular Function (MF) terms. Additionally, the enrichment of DMPs in six classic histone modifications was assessed using ChIP-seq data from 18 blood cell type-related epigenomes, sourced from the Roadmap Epigenomics Consortium. Chromatin state annotations for these same samples were derived using the NIH Roadmap’s ChromHMM expanded 18-state model. To evaluate DMP enrichment within transcription factor binding sites (TFBS), data from human meta-clusters available in the Gene Transcription Regulation Database (GTRD) were employed (v18.06). Statistical significance of the observed enrichments was determined using one-sided Fisher’s exact tests (*adjusted p-value* < 0.05), comparing the overlap of DMPs with the relevant dataset against a background set of 647,153 filtered probes from the HumanMethylationEPICv2 array.

## RESULTS

### *De novo* pathogenic variants in 3 snRNA genes in patients with NDD

The detailed phenotypic characterization of patients enrolled in the Spanish URD Programs and comprehensive genome variant analyses enabled the systematic investigation of potentially pathogenic variants affecting distinct functional domains of candidate snRNA genes and their association with NDDs. We identified potentially pathogenic heterozygous variants in three of the four snRNA genes of the major spliceosome that have similarly constrained domains and are expressed in brain^6^, *RNU4-2*, *RNU2-2*, and *RNU5B-1* (**Table 1 & Figure 1A,1B**), in 38 patients with previously undiagnosed NDD. No candidate variants were detected in *RNU4-1*.

**Figure 1.**
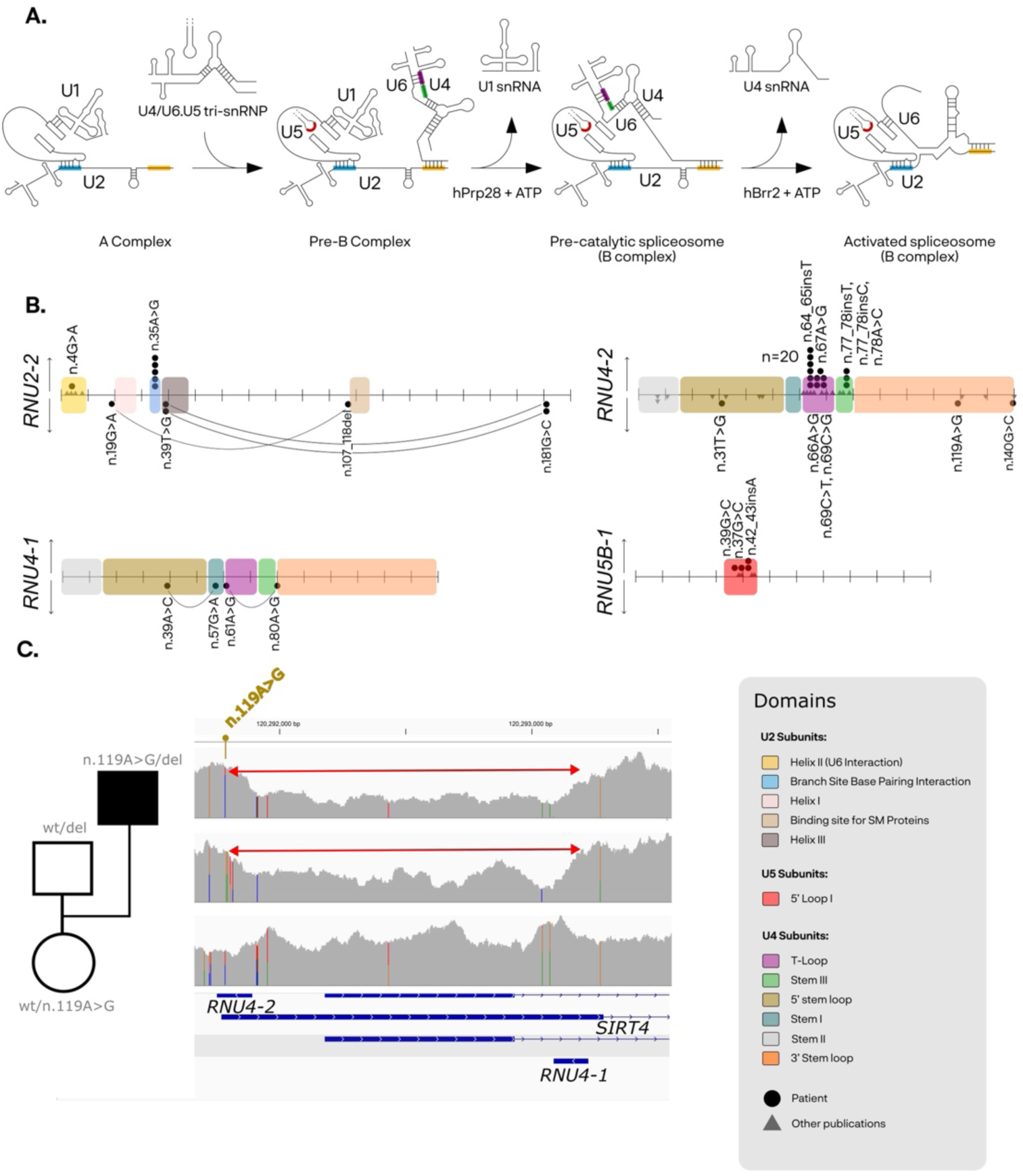
Schematic representation of pathogenic variants detected in snRNA genes in NDD patients from the Spanish URD Programs. **A.** Schematic representation of the major spliceosome complex. Illustrating the dynamic transitions through different intermediate stages: A complex, pre-B complex, pre-catalytic (B complex) and activated spliceosome (B complex). Highlighted in colour are the domains of the snRNA genes (U2, U4, and U5) where the identified mutations are located. Adapted ^52^ **B.** Variant distribution across snRNA genes in our cohort. The upper panel shows de novo variants, while the lower panel displays biallelic variants. Lines connect variants found in the same individual, representing combinatorial configurations. Colors indicate the structural domain in which each variant is located. Circles represent individuals from our cohort, whereas triangles correspond to previously reported variants from published studies. **C.** Visualization of the deletion identified in patient U4-p32, which spans both RNU4-2 and RNU4-1 Genome browser views of the patient and both parents are shown for comparison. The yellow arrow marks the single nucleotide variant (SNV) located within the affected region. The red arrow marks the deletion.

**Table 1.** Pathogenic/likely pathogenic variants in snRNA genes in patients with NDD from the Spanish URD Programs. The table includes variant-related information, such as HGVS nomenclature, chromosomal location (GRCh38) and altered gene domain. It is also indicated the total number of variant carriers among Spanish undiagnosed NDDs, inheritance pattern and genotypes. Except for n.66A>G, which has an allele count (AC) of 1, all variants have ACs of 0 in gnomAD v4.1. Variants n.69G>C and n.77_78insC have not been previously described. *Previously reported **Patient is also a carrier of a genomic duplication encompassing HNRNPH1 and RUFY1, reported as likely pathogenic ^43^ (Case 3). ?: unknown; non-paternal, maternal sample unavailable. MZT: monozygotic twins.

| Gene | Variant | Position (GRCh38) | Domain | AC gnomADv4 | Proband | Dominant de novo | Recessive biallelic | Genotype |
| --- | --- | --- | --- | --- | --- | --- | --- | --- |
| RNU2-2 | n.4G>A | chr11:62841622 | U6 binding domain | 0 | 1* | 1 | - | het |
|  | n.35A>G | chr11:62841703 | Branch site recognition domain | 0 | 5* | 5 | - | het |
| RNU4-2 | n.64_65insT | chr12:120291839 | T-loop (quasi-pseudoknot) | 0 | 20* | 19+? | - | het |
|  | n.66A>G | chr12:120291838 | T-loop (quasi-pseudoknot) | 1 | 2 (MZT) | 2 | - | het |
|  | n.67A>G | chr12:120291837 | T-loop (quasi-pseudoknot) | 0 | 1 | ? | - | het |
|  | n.69C>T | chr12:120291835 | T-loop (RBM42 interaction) | 0 | 1* | 1 | - | het |
|  | n.69C>G | chr12:120291835 | T-loop (RBM42 interaction) | 0 | 1 | 1 | - | het |
|  | n.77_78insC | chr12:120291826 | Stem III | 0 | 1 | 1 | - | het |
|  | n.77_78insT | chr12:120291826 | Stem III | 0 | 1 | 1 | - | het |
|  | n.78A>C | chr12:120291826 | Stem III | 0 | 1 | 1 | - | het |
| RNU5B-1 | n.37G>C | chr15:65304713 | 5' loop I | 0 | 1 | 1 | - | het |
|  | n.39C>G | chr15:65304715 | 5' loop I | 0 | 1 | 1 | - | het |
|  | n.42_43insA | chr15:65304718 | 5' loop I | 0 | 2 | 2 | - | het |
| Subtotal |  |  |  |  | 38 |  |  |  |
| RNU2-2 | n.19G>A | chr11:62841791 | Branch site recognition domain | 1 | 1 | - | pat | comp het |
|  | n.107_118del | chr11:62841679-62841690 | Binding site SM proteins | 4 |  | - | mat |  |
|  | n.39T>G | chr11:62841771 | Stem Ia | 0 | 2 (Sibs) | - | pat | comp het |
|  | n.181G>C | chr11:62841629 | - | 6 |  | - | mat |  |
| RNU4-1 | n.39A>C | chr12:120293199 | 5' Stem loop | 138 | 1 | - | pat | comp het |
|  | n.57G>A | chr12:120293181 | Stem I | 3 |  | - | mat |  |
|  | n.61A>G | chr12:120293177 | Stem I | 35 | 1 | - | pat | comp het |
|  | n.80A>G | chr12:120293158 | Stem III | 12 |  | - | mat |  |
| RNU4-2 | n.31T>G | chr12:120291873 | 5' stem loop | 0 | 1** | - | mat & pat | hom |
|  | n.140G>C | chr12:120291764 | 3' stem loop | 1 | 1 | - | mat & pat | hom |
|  | n.119A>G | chr12:120291785 | 3' Stem loop | 8 | 1 | - | mat | comp het |
|  | 1.778kdel | chr12:120291763-120293127 | Entire U4-1 & U4-2 genes | 0 |  | - | pat |  |
| Subtotal |  |  |  |  | 8 |  |  |  |

A total of 28 individuals had heterozygous pathogenic variants in the *RNU4-2* gene (**Figures S1, S2A**). All probands, including a monozygotic twin pair, had unaffected parents. The recurrent single nucleotide insertion variant n.64_65insT was found in 20 patients (70%), confirming it is a mutational hotspot. Other variants previously reported in other patients included n.66A>G, found in the monozygotic twins, n.67A>G, n.69C>T, and n.77_78insT. Furthermore, we identified two novel variants, n.69C>G and n.77_78insC in single cases. All these variants lie in the central region of the U4/U6 snRNA duplex and are predicted to affect intermolecular interactions. *De novo* occurrence was confirmed, either by genome trio analysis and/or Sanger sequencing, in all cases but two, both conceived by in vitro fertilization using anonymous egg donors.

We also identified 6 individuals with *de novo* pathogenic variants in *RNU2-2* (n.35A>G in 5 patients and n.4G>A in 1 patient), who have been already reported (**Figure S2B**)^5^.

Among the additional screened snRNA genes under evolutionary constraint, we found 4 unrelated affected individuals with heterozygous variants in *RNU5B-1* that were also confirmed to occur *de novo*. The identified variants (n.42_43insA in two cases, n.37G>C and n.39C>G in one case each) clustered within the highly conserved 5′ loop I domain of the gene (**Figures S1, S3**). All 4 patients with *de novo RNU5-B1* variants presented with global developmental delay, motor dysfunction, and craniofacial dysmorphisms, with overlapping clinical profiles to those of the other RNU-related syndromes^7,8^.

No evidence of rare variant enrichment and no single *de novo* variant was found in other U5 family genes (*RNU5A-1*, *RNU5D-1, RNU5E-*1, *RNU5F-1*). For a detailed description of all the identified variants, see **Tables S4, S5**.

### Autosomal recessive NDD caused by biallelic snRNA gene variants

We then explored whether snRNA genes can also contribute to disease via autosomal recessive inheritance, searching for biallelic variants across the cohort. A set of 19 candidate snRNA genes were selected based on their expression profiles in disease-relevant tissues (brain) using GTEx data (**Table S6 & Figure S4**). We identified rare biallelic variants in 8 probands involving 3 of those snRNA genes, *RNU2-2*, *RNU4-1*, and *RNU4-2* (**Figure 1A 1B, Table 1, Table S7**).

Three probands with NDDs carried biallelic variants in *RNU4-2*. One of them, case U4-p32, was compound heterozygous for a maternally inherited variant (n.119A>G) and a paternally inherited 1364 bp deletion. The n.119A>G variant has been reported in other individuals with neurodevelopmental phenotypes, both in homozygous^16^ and compound heterozygous patterns. The heterozygous 1364 bp deletion leads to the fusion of the *RNU4-1* and *RNU4-2* genes, so it seems to have been mediated by unequal recombination between these two highly similar loci (**Figure 1C**). Case U4-p32 had a brother with a similar NDD who died at young age, and a healthy sister; however, no DNA sample from the affected sibling is available to determine whether he carried the variants. No other rare structural variants were identified within a ±2000 base pair window around the studied genes. A second proband, case U4-p31, was homozygous for the n.31T>G variant, located in a functionally constrained genomic region depleted of variation in population databases. This patient also carries a copy number gain disrupting the genes *HNRNPH1* and *RUFY1* that has been previously reported as likely pathogenic^43^. The n.31T>G variant has also been reported in homozygosity in an individual with mild intellectual disability from the UK Biobank^16^. The third patient, case U4-p33, was homozygous for a n.140G>C substitution, located within the 3′ stem-loop structure of the *RNU4-2* RNA. Interestingly, a different substitution in the same nucleotide position has also been reported in homozygosity in affected siblings with NDD (chr12: 120291764C>T, n.140G>A)^16^.

Three probands, case U2-p7 and two affected siblings (cases U2-p8 and U2-p8s), carried compound heterozygous variants in *RNU2-2* (**Table 1, Table S7**). All three individuals exhibited neurodevelopmental delay, and two of them a drug-resistant epileptic encephalopathy (U2-p7 and U2-p8s). Notably, the identified mutations do not cluster within any known annotated functional domain of the gene (**Figure 1B**). Overall, these 5 individuals with biallelic *RNU4-2* and *RNU2-2* variants showed overlapping phenotypes with those reported in carriers of *de novo* variants in the same genes, reinforcing their potential role in autosomal recessive forms of snRNA-associated disorders and extending previous evidence implicating *RNU4-2* in recessive neurodevelopmental syndromes to suggest similar contributions from other snRNA loci.

Two individuals were compound heterozygous for rare variants at *RNU4-1* (**Table 1, Table S7**). This gene, which differs from *RNU4-2* by only four nucleotide changes (97.2% sequence identity) and shows a similar expression pattern, has not yet been associated with disease by *de novo* variants. The biallelic variants found are located near conserved structural motifs where pathogenic *de novo* variants have been identified in *RNU4-2*, next to Stem III (n.80A>G) or within the T-loop motif (n.57G>A; n.61A>G) regions. These two cases with biallelic *RNU4-1* variants displayed a severe NDD with global developmental delay and dysmorphic features, along with spastic tetraparesis in one individual (case U41-p1). Brain imaging revealed polymicrogyria in U41-p1 and hypomyelination in U41-p2, highlighting phenotypic variability that complicates interpretation of the gene’s involvement in NDD.

### Possible digenic inheritance

We also explored whether digenic inheritance involving variants in two different snRNA subunits may underlie disease in a subset of cases with NDD. We investigated whether combinations of damaging variants in at least two of the same 19 candidate snRNA genes were present in patients more often than would be expected by chance by comparing NDD cases and controls (parents and individuals with non-NDD phenotypes). We found a modest but significant enrichment of digenic variant combinations in NDD individuals compared to controls (OR=2.08, p-value=0.04) (**Figure S5**), suggesting that di-oligogenic mechanisms may also contribute to disease in some cases. While not conclusive, this points to a more complex genetic architecture that warrants further investigation through functional studies and larger cohorts.

### snRNA gene variants can associate different phenotypes

The n.7G>A variant at *RNU4-2* was previously reported in compound heterozygosity with n.11A>C in an individual with an undiagnosed NDD^16^. In our cohort, we identified the n.7G>A variant in *trans* with a second variant, n.18_19insA, in the proband of a large family with vertical transmission of adolescent-onset rod-cone dystrophy but no other neurodevelopmental problems^44^. This indel, located between stem II and the U4 5’ stem-loop, has also been observed in other unsolved cases of retinitis. All affected individuals with rod-cone dystrophy in this family (**Figure S6**) carried the n.18_19insA variant in a monoallelic state, except for one branch in which it co-occurred with the n.7G>A variant. The n.7G>A variant was present in the unaffected father and a brother. The absence of phenotypic differences between monoallelic and biallelic carriers supports a dominant mode of inheritance for the insertion. The n.7G>A variant appears to have no effect on the ocular phenotype, nor does it cause neurological symptoms when present in a single allele, in this context is therefore considered benign. Its clinical relevance has only been described in compound heterozygosity with other *RNU4-2* variants probably located within the same stem II region contributing to NDD, but not to ocular disease^16^.

A single individual with congenital horizontal nystagmus and no other neurodevelopmental problems carried biallelic variants in *RNU1-27P* (**Table S7**), one of the snRNAs with stronger expression in human tissues, including brain, liver, and pancreas (**Figure S3**). However, no functional annotations or prior disease associations are currently available for this gene, so the identification of additional patients and/or further research to define the functional consequences of this variant are needed to establish the potential role of *RNU1-27P* in pathology.

### Predicted snRNA secondary structures

While *de novo* dominant variants in *RNU4-2* and *RNU2-2* are though to disrupt critical motifs involved in branch point recognition and spliceosomal assembly, the pathogenic mechanisms of *RNU5B1* and biallelic variants are still unknown. Predicted consequences of the detected *RNU5B-1* variants on RNA secondary structure were modelled using RNAfold^28^. The indel (n.42_43insA) variant creates an enlarged loop sifted a few nucleotides towards the 5’ end in the predicted RNA folding compared to the wild type, leaving room for a possible structural effect of this mutation, whereas the other two SNVs maintain the overall structural conformation (**Figure S7**). These results suggest that *RNU5B-1* variants may also exert their pathogenic effect through structural disruption and/or impairing specific nucleotide interactions through loop I, essential for exon recognition and splicing fidelity. This similar mechanism for most de novo variants in the different snRNA subunits suggests a dominant negative effect. For biallelic variants, RNAfold predictions consistently revealed structural alterations, except in cases where the variant was located within a loop region, likely affecting specific interactions rather than the overall RNA conformation (**Figure S7**).

### Deep phenotypic profiling of snRNA related NDDs

To define the phenotypic variability and gene-specific effects of snRNA-associated disorders, we performed an in-depth clinical characterization of the 38 patients with pathogenic *de novo* variants in *RNU2-2*, *RNU4-2* or *RNU5B-1*. These analyses yielded 200 unique HPO terms (**Table S8**). Reverse phenotyping was then performed to ensure consistent and complete data. Unsupervised clustering based on binary presence/absence of HPO terms revealed three principal phenotypic groups. Two clusters were dominated by *RNU4-2* patients, each including one *RNU5B-1* individual with the n.42_43insA variant, suggesting phenotypic overlap for this specific mutation. A third, more heterogeneous cluster comprised all *RNU2-2* cases alongside the remaining two *RNU5B-1* individuals. Within this cluster, *RNU2-2* patients displayed a highly homogeneous clinical profile, while *RNU5B-1* cases showed more variability. Notably, these clustering patterns did not correlate with age, reinforcing the role of genotype as the main phenotypic driver (**Figure S8**).

Despite gene-related variability, neurodevelopmental features shared by all patients included global developmental delay, intellectual disability, motor delay, and speech and language impairment. Musculoskeletal findings, including generalized hypotonia and growth abnormalities such as failure to thrive and short stature, were also frequent. Craniofacial dysmorphisms were broadly present, though their specific characteristics varied between genes (**Figure 2A**).

**Figure 2.**
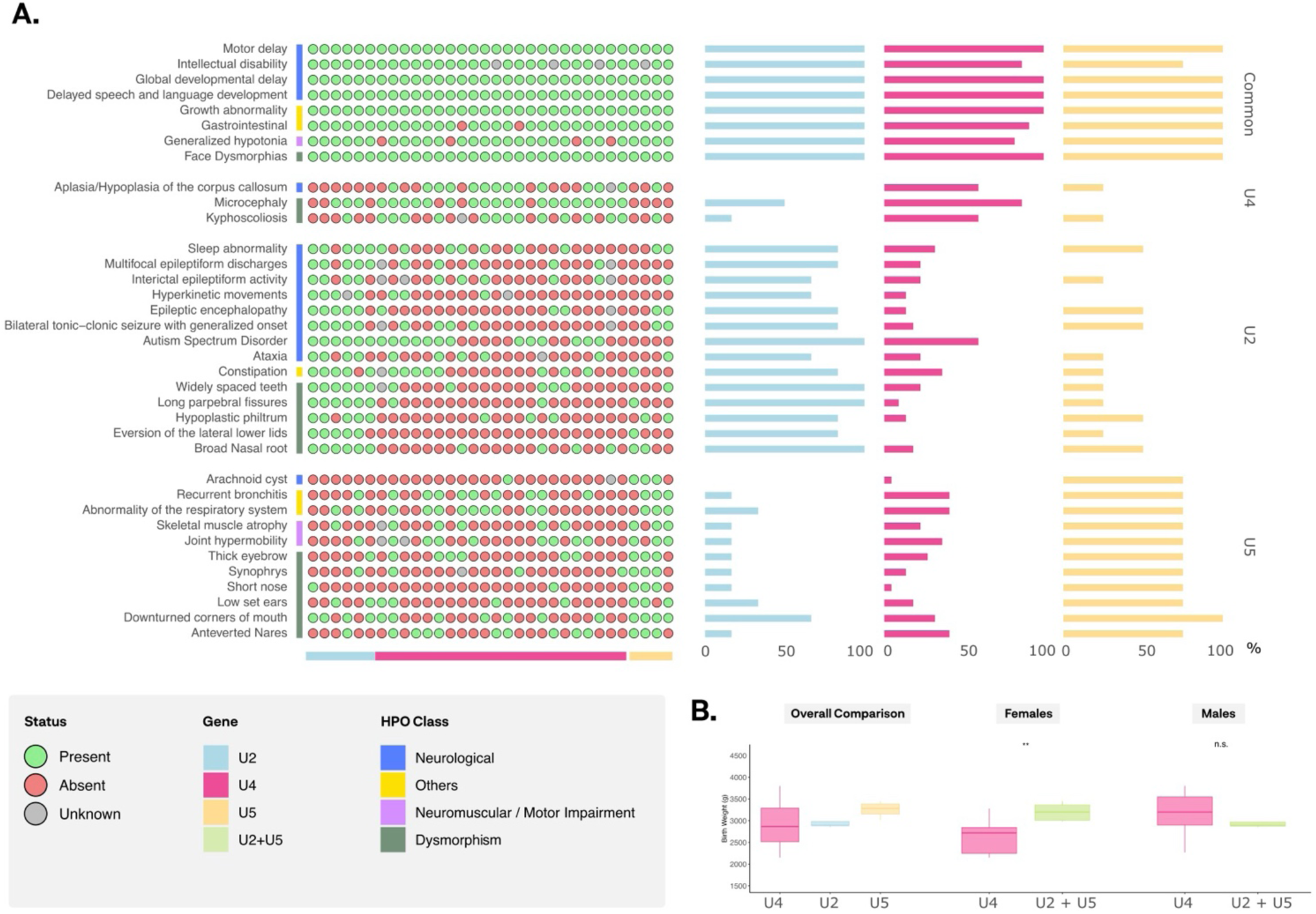
Phenotypic spectrum and birth weight di8erences in individuals with de novo snRNA variants. **A.** Left panel: Human Phenotype Ontology (HPO) representation of the most common phenotypic features observed in patients with mutations in each snRNA gene (U2, U4, and U5), along with the specific HPO terms unique to each group. Phenotypes are categorized into major classes: neurological (blue), neuromuscular / motor impairment (purple), dysmorphic features (green), and other (yellow). Right panel: Overall HPO frequency distribution across each group (U2, U4, and U5). **B.** Birth weight distribution stratified by sex. After excluding preterm cases, Individuals with RNU4-2 variants were compared to those with variants in RNU2-2 or RNU5B-1 (grouped as RNU2-2 + RNU5B-1), using two-sample t-tests for each comparison. P < 0.05 (*); not significant (n.s.).

Beyond these shared clinical manifestations, some gene-related distinctive features emerged. *RNU4-2* was closely associated with craniofacial and brain anomalies, particularly microcephaly, hypotelorism, and corpus callosum hypoplasia, as well as kyphoscoliosis. *RNU2-2* was strongly linked to severe epileptic encephalopathy, often accompanied by developmental regression. Craniofacial features such as broad nasal root, widely spaced teeth, and long palpebral fissures were also consistently observed. In contrast, *RNU5B-1* showed a broader phenotypic spectrum. Case reports for each *RNU5-B1* patient are also available (**Supplementary Material**). While neurodevelopmental delay was common, other traits, such as epilepsy, behavioral symptoms, and craniofacial anomalies (e.g., low-set ears, thick eyebrows), varied between individuals (**Figure 2A**).

To explore intragenic variation, we assessed whether mutation location in *RNU4-2* could influence the phenotype as previously described^7^. Pathogenic variants clustered mainly in the T-loop (25 patients) and stem III (3 patients). While no consistent domain-specific phenotypic patterns were identified, likely due to the limited sample size for stem III, differences were observed in the severity of intellectual disability, global developmental delay, speech impairment, and seizure phenotypes. Overall, individuals with T-loop variants presented more severe clinical features than those with Stem III variants, who exhibited a milder phenotype.

A prenatal phenotype was notorious in patients with *RNU4-2* variants, particularly females, who exhibited significantly increased rates of prematurity, intrauterine growth restriction (IUGR) and lower birth weights. Prematurity was observed in 4/13 (30%) females and 1/12 males with *RNU4-2* variants, while all patients with *RNU2-2* and *RNU5B-1* variants were born at term. Although one-way ANOVA did not reveal significant differences across genotypes overall, sex-stratified analyses highlighted a significant reduction in birth weight among females with *RNU4-2* variants compared to females harboring other variants (*p=0.0068*) after excluding preterm cases (**Figure 2B**). We also observed a notable enrichment of IUGR among *RNU4-2* cases—a feature not reported in *RNU2-2* or *RNU5B-1* individuals. Nearly half (48%) of *RNU4-2* patients presented with prenatal IUGR, with a marked female predominance (70% of females versus 33% of males). Additionally, mild ventriculomegaly was detected on prenatal ultrasound scans in several *RNU4-2* cases (three females and two males). One case involved a twin pregnancy conceived via egg donation, where the demise of one twin during the first trimester was detected along with the presence of ventriculomegaly in the surviving fetus (**Table S9**). These findings emphasize the importance of including targeted testing for *RNU4-2* variants in prenatal setting, in pregnancies with unexplained IUGR and other anomalies.

### Distinct craniofacial features and facial mask for *RNU4-2* syndrome

Facial pictures of the patients are available in **Figure S9**. To explore gene-specific craniofacial patterns in snRNA-associated disorders, we analyzed frontal facial images from individuals with pathogenic variants in *RNU2-2* (n=10), *RNU4-2* (n=15) and *RNU5B-1* (n=4), along with 31 unaffected controls, using DeepGestalt and GestaltMatcher algorithms (Face2Gene Research platform, FDNA Inc., USA). Based on similarity rankings generated by DeepGestalt, Coffin-Siris syndrome (*CSS1;* OMIM# 135900) and *MRD5*-associated disorder (OMIM#612621) were identified as the closest phenotypic matches to *RNU4-2* and *RNU2-2*, respectively, and were thus included as reference groups.

Multiclass classification across all cohorts yielded a mean accuracy of 53.3%, exceeding random expectation (39.2%) and suggesting partially distinct facial profiles. Binary comparisons revealed that *RNU4-2* was clearly distinguishable from both unaffected controls (AUC=0.948, *p=0.003*) and *RNU2-2* (AUC=0.921, *p=0.035*), indicating a consistent and well-defined facial signature. Conversely, *RNU2-2* could not be robustly separated from controls (AUC=0.741, *p=0.121*), reflecting a milder and more variable craniofacial presentation (**Figure 3A, 3B**).

**Figure 3.**
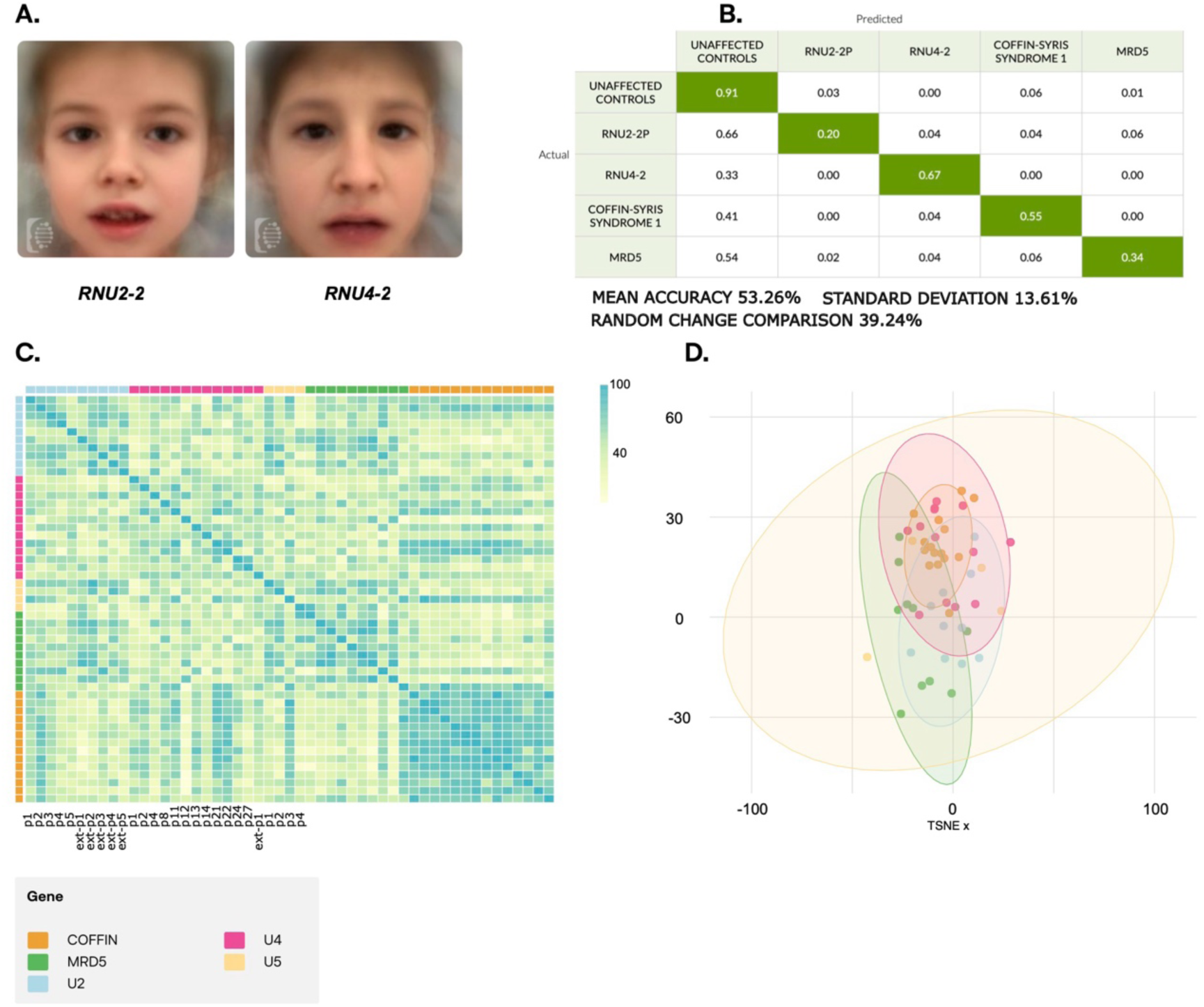
Facial analysis of individuals with de novo snRNA variants. **A.** Composite facial masks generated using the FDNA system for individuals with U4, U2 variants, and unaZected individuals. **B.** Confusion matrix displaying True Positive (TP) values on the diagonal, while oZ-diagonal values represent False Positive (FP) and False Negative (FN) rates. **C.** Pairwise Comparison Matrix (PCM), showing facial similarity scores among individuals in the cohort. Higher similarity in phenotypic features is indicated in blue, with a value of 100 representing identical comparisons. **D.** t-SNE visualization illustrating the clustering of facial phenotypes across the cohort.

GestaltMatcher analysis showed that individuals with *RNU4-2* and *RNU2-2* variants consistently clustered with other cases from the same gene group and with their respective confusion syndromes, *CSS1* for *RNU4-2* and *MRD5* for *RNU2-2*, reinforcing the gene-specific facial signatures. In contrast, individuals with *RNU5B-1* variants were more dispersed across the facial phenotype space, showing no consistent clustering pattern (**Figure 3C, 3D**).

These results suggest that ReNU syndrome is associated with a quite consistent and distinguishable facial phenotype, whereas the facial features of cases with *RNU2-2* variants largely overlap with those of unaffected individuals. The broader distribution observed in *RNU5B-1* cases further underscores the phenotypic heterogeneity of this still small group, aligning with the clinical variability highlighted by HPO-based analyses.

### *RNU4-2*-specific episignature

We also obtained methylation array profiling to explore epigenetic alterations in patients with snRNA gene variants. In individuals with *de novo RNU4-2* mutations, we identified a distinct episignature, particularly among those carrying variants in the T-loop domain. A total of 182 probes were selected through the episignature detection pipeline (**Table S10**). This signature was validated by hierarchical clustering and multidimensional scaling (**Figure 4A, 4B**), and its robustness was confirmed via leave-one-out cross-validation (LOOCV) (**Figure S10A**). A support vector machine (SVM) classifier built on these probes correctly identified 4 out of 5 independent validation samples (**Figure S10B**). The only misclassified sample carried the n.77_78insG variant, located in a different domain, consistent with the previous results^7^. Although there was no direct overlap between the top-ranked probes from our analysis and those in another published episignature, we intersected the probe list with the statistics generated by our own limma analysis. For the great majority of these probes, the Δβ direction (hypo-vs hypermethylation) was concordant (**Figure S10C**), and the Pearson correlation of Δβ values reached 0.78, further confirming that our data recapitulate similar biological signals and supporting the robustness of the episignature^7^.

**Figure 4.**
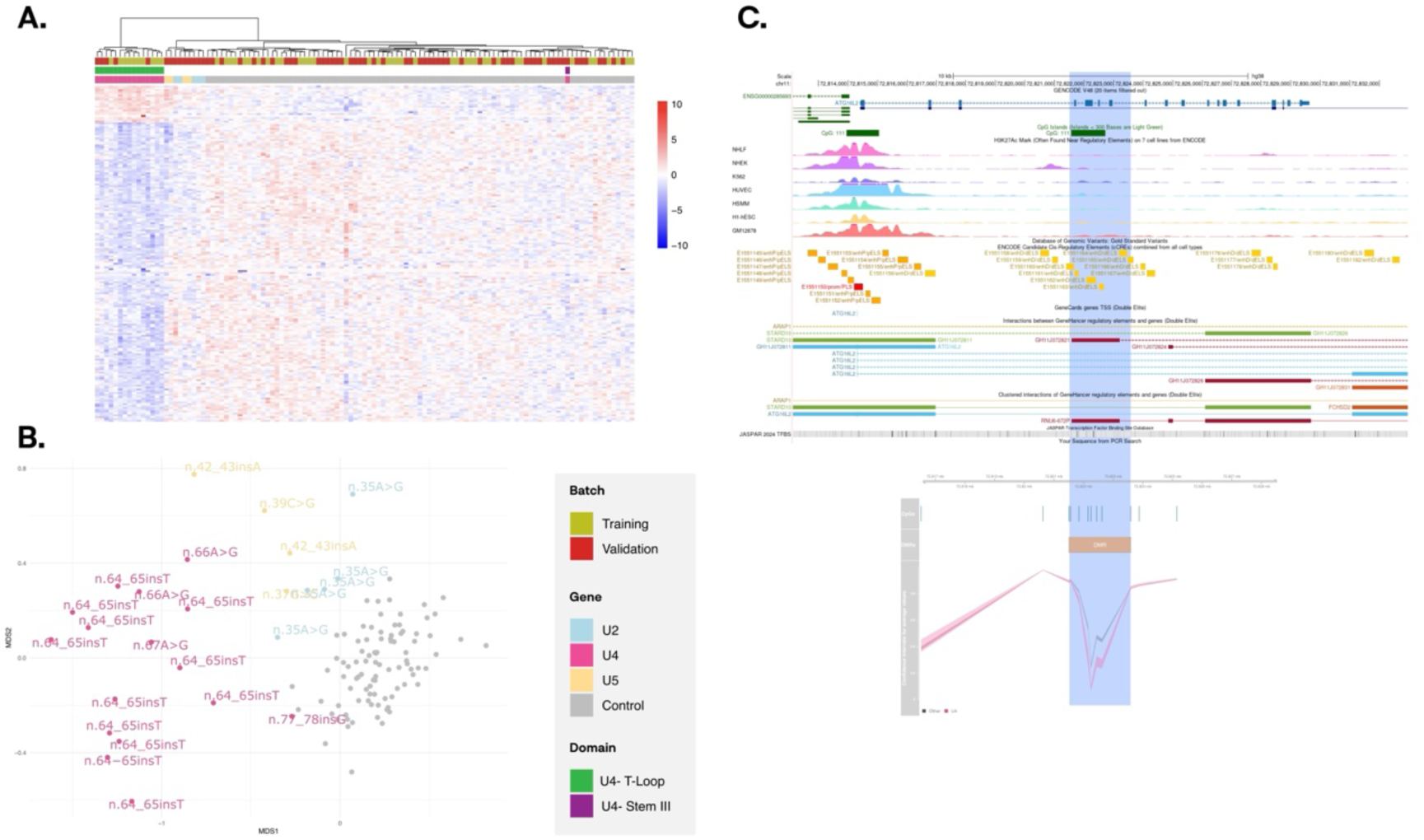
**DNA methylation analyses in individuals with de novo RNU4-2 variants**. **A.** Hierarchical clustering analysis using U4-specific probes. Rows correspond to selected probes, while columns represent individual samples. Methylation levels range from blue (hypomethylated) to red (hypermethylated). U2, U4, U5, and control groups are labelled in blue, pink, yellow, and grey, respectively. Samples are also categorized into training (red) and validation (green) cohorts. **B.** Multidimensional Scaling (MDS) plot of selected probes, with U2, U4, U5, and control groups represented in blue, pink, yellow, and grey, respectively. **C.** DiZerentially methylated region (DMR) identified in the promoter of RNU6-672P. Upper panel: UCSC Genome Browser screenshot displaying the genomic context of the DMR, including annotated genes (blue), CpG islands (green), HЗK27Ac marks, ENCODE cis regulatory elements (transcription start sites (TSS; red), enhancers (E; yellow), and weak enhancers (WE; pale yellow)), GeneHancer annotations and JASPAR Transcription factor binding site. Lower panel: plot showing methylation levels (mean ± confidence intervals) for each CpG probe within the DMR in U4 cases (pink) and others (gray).

For *RNU2-2* and *RNU5B-1*, although several differentially methylated probes (DMPs) were detected, robust episignatures could not be defined, most likely due to the limited number of available samples. LOOCV revealed variability across iterations, highlighting the need for larger cohorts (**Figure S11**).

### Consequences of aberrant methylation in ReNU patients

Given that the main pathogenic mechanism of snRNA related NDDs is thought to be the quantitatively aberrant splicing of relevant target genes important for neurodevelopment, as reflected in transcriptomic analyses, DNA methylation changes can also inform about downstream regulatory effects directly or indirectly involved in pathogenicity. To explore the biological implications of these methylation changes, we performed functional enrichment analysis. No Gene Ontology (GO) terms reached statistical significance after FDR correction. However, histone mark enrichment analysis revealed that differentially methylated probes (DMPs) were significantly enriched in enhancer regions marked by H3K4me1, as well as in repressive heterochromatin regions marked by H3K27me3 and H3K9me3 across nearly all analyzed blood-derived epigenomes (**Figure S10D**). Further inspection of chromatin states where these histone marks co-occurred indicated notable enrichment in gene regions encoding Zinc Finger proteins and in Polycomb-repressed chromatin (**Figure S10E**). In addition, transcription factor binding site (TFBS) analysis using the GTRD database revealed highly significant enrichment for SMC1A, SMC3 and RAD21 (**Figure S10F**), core components of the cohesin complex. All three proteins have been implicated in Cornelia de Lange syndrome (OMIM#122470)^45^, a NDD that shares some phenotypic traits with patients carrying RNU gene variants.

In addition, we investigated differentially methylated regions (DMRs) within the subset of *RNU4-2* patients who clustered within the identified episignature. We detected eight significant DMRs, the majority of which were hypomethylated (**Table S11**). Some DMRs identified overlap with genes highly expressed in the brain that could influence crucial aspects of neurodevelopment and synaptic function (**Figure S12).** *ATG16L2* is involved in autophagy, which is essential for neurogenesis and synaptic plasticity. *LRFN1* (SALM2) is a synaptic adhesion molecule playing a role in neural circuit development. Additionally, *CALN1*, highly expressed in cerebellum, is involved in calcium homeostasis, a key factor for neuronal communication. Interestingly, one hypomethylated DMR overlapped the promoter of *RNU6-672P*, a pseudogene with 89% sequence identity to *RNU6-1*, a core component of the U4/U6 spliceosomal complex (**Figure 4C**). This epigenetic alteration might be an additional target effect or might reflect a compensatory regulatory mechanism aimed at modulating transcription in response to abnormal spliceosome and U4-related splicing dysfunction.

## DISCUSSION

Our data further support the role of *de novo* dominant variants in several snRNA genes, *RNU4-2*, *RNU2-2*, and *RNU5B-1*, as causative of NDDs, consistent with other recent studies^4–8^. Disease-causing mutations clustered in highly conserved functional domains: the T-loop and stem III in *RNU4-2*; the branch point recognition motif and helix II in *RNU2-2*; and loop I in *RNU5B-1*. These structural disruptions lead to distinct neurodevelopmental phenotypes. Mild alterations in alternative splicing have been observed in blood-derived RNA from individuals with pathogenic *RNU4-2* variants, but similar effects remain undocumented for *RNU2-2* and *RNU5B-1*. The subtlety or absence of detectable splicing aberrations in peripheral tissues might be related to a brain-specific and/or developmental pathophysiology, underscoring the need for future studies, including animal models and RNA-seq in neural tissue, to better elucidate the functional consequences in disease-relevant cell types and developmental stages.

While most identified cases follow the *de novo* dominant pattern, we also detected individuals harboring rare homozygous or compound heterozygous inherited variants in snRNA genes. Heterozygous carriers are healthy with no phenotype. Most snRNA variants associated with recessive conditions lie outside canonical functional domains, suggesting alternative pathogenic mechanisms. Two recent studies^16,17^ reported 32 individuals with NDDs due to recessive *RNU4-2* variants. Three of the reported variants (n.119A>G, n.31T>G, and n.7G>A) have also been found in either homozygosity or compound heterozygosity in unrelated probands of our cohort. Notably, n.119A>G was found in *trans* with a *RNU4-2* gene deletion likely generated by recombination between the *RNU4-2* and *RNU4-1* genes. To our knowledge, this is the first described copy number alteration affecting RNU genes as a mechanism of biallelic disruption. The data indicate that *RNU4-2* is haplosufficient and biallelic loss of function variants are required for disease, while a dominant negative effect may be responsible for the dominant / *de novo* inheritance of the disease, as commonly found in other protein-coding genes^46^.

The *RNU4-2* variant n.7G>A, previously associated with NDD in a compound heterozygous state^16^, was identified in *trans* with n.18_19insA in three siblings of a family with multiple affected members affected with a retinal dystrophy of similar severity and no neurodevelopmental issues^44^. Only the indel variant (n.18_19insA) segregated with the dominant phenotype in the family. Therefore, variant n.7G>A seems to be benign in heterozygous state and in combination in *trans* with n.18_19insA in this family. These findings underscore the importance of careful variant interpretation in *RNU4-2*, particularly given the gene’s dual involvement in ocular and neurological phenotypes depending on variants, combinations and zygosity.

Additionally, we identified three individuals carrying biallelic variants in *RNU4-2,* three in *RNU2-2*, and two in *RNA4-1*, indicating that variants in these genes can also associate autosomal recessive NDDs. The phenotypes observed in individuals with biallelic variants in *RNU2-2* and *RNU4-2* overlapped with those reported in *de novo* variant carriers of the same genes, reinforcing their pathogenic relevance. However, detailed phenotypic characterization may uncover subtle distinctions between inheritance models, and functional studies are needed to clarify the underlying mechanisms. These findings also raise the possibility that snRNA-associated disorders may, in some cases, result from digenic or oligogenic inheritance, involving combinations of pathogenic variants across different subunits of the major spliceosome complex. Indeed, we found a significant enrichment of digenic rare variant combinations in NDD individuals compared to controls (OR=2.08), which should be further investigated in other cohorts.

Deep phenotypic profiling of individuals with *de novo* variants, including structured clinical coding through HPO terms and automated facial analysis, enabled us to delineate gene-specific features. *RNU4-2* is associated with craniofacial and structural brain abnormalities such as microcephaly, hypotelorism and corpus callosum hypoplasia. Notably, a distinct facial gestalt was identified in individuals with pathogenic variants in *RNU4-2*, which effectively distinguished affected individuals from both controls and *RNU2-2* cases. This recognizable facial “mask” reinforces the gene-specific nature of the syndrome and may serve as a useful diagnostic clue, particularly in contexts where access to genetic testing is limited or delayed. In contrast, *RNU2-2* variants were linked to early-onset epilepsy and developmental regression, consistent with a more severe neurodevelopmental phenotype^5^, but showed milder and less distinctive facial features, limiting their utility for automated recognition. *RNU5B-1* cases exhibited broader clinical variability, possibly due to a limited sample size, although other studies also suggest more heterogeneous clinical expressivity for this gene ^7,8^.

A prenatal phenotype, with IUGR leading to low birth weight and mild brain ventriculomegaly on ultrasounds, was present in individuals with *de novo RNU4-2* variants, mainly in females, but not observed in *RNU2-2* or *RNU5B-1* variant carriers. Thus, *RNU4-2* disruption may affect prenatal development beyond the nervous system, with a potential sex-specific vulnerability in female fetuses. IUGR has also been observed in fetuses with *RNU4ATAC* mutations, ranging from mild to severe^47,48^. Based on our findings, prenatal abnormality could raise clinical suspicion of *RNU4-2* mutations, supporting the need for targeted prenatal genetic testing. In addition, *RNU4-2* should be considered in cases of unexplained prenatal IUGR, particularly in females, with negative QF-PCR and prenatal exome results, as has previously been recommended for other disorders of the spliceosome^49^.

We also observed that variant location within *RNU4-2* appears to influence clinical severity. Mutations affecting the T-loop domain were generally associated with more severe phenotypes, including profound intellectual disability and seizures, whereas variants in stem III were linked to milder presentations. Although the number of cases per domain remains limited, this correlation aligns with previous reports^7^ and is further supported by epigenetic data.

Specifically, we identified a reproducible blood-derived episignature associated with T-loop mutations, which was consistently and accurately classified using support vector machines (SVM), in line with prior findings. Despite limited probe overlap with published episignatures, the robustness of the classification suggests the existence of a domain-specific methylation pattern. These results underscore the utility of methylation profiling as a functional biomarker. It remains to be explored whether biallelic variants, currently underrepresented in methylation studies, give rise to distinct or more variable episignatures, which could provide deeper insight into their pathogenic mechanisms. Beyond classification, our epigenetic analyses also provide biological insight into the downstream effects of these mutations. Functional enrichment analysis revealed significant enrichment for binding sites of cohesin complex proteins known to be involved in Cornelia de Lange syndrome^45^, a NDD with overlapping phenotypic features. Moreover, recent proteomic and functional studies have demonstrated that cohesin interacts directly with multiple splicing factors and RNA-binding proteins, including components of the U4/U6/U5 tri-snRNP complex^50^, and actively regulates alternative splicing in both physiological and pathological contexts^51^. Together, these findings suggest a mechanistic link between the methylation changes observed in our cohort, cohesin complex activity, and spliceosomal function, offering a plausible molecular basis for the neurodevelopmental phenotype. Additionally, we detected differentially methylated regions near neurodevelopmental genes and within the promoter of the *RNU6-672P*. pseudogene, pointing to a possible epigenetic mechanism regulating U6 snRNA. This may impact U4/U6 complex assembly and spliceosomal function, providing a hypothesis for further investigations.

The integrated Spanish programs for URD, IMPaCT-Genómica, URD-Cat, and ENoD-CIBERER, have demonstrated substantial clinical utility. Their commitment to equitable access has enabled hospitals across Spain to benefit from standardized protocols and advanced diagnostic tools including WGS to patients with URDs. By analyzing WGS from over 1,708 probands with neurodevelopmental disorders (NDDs) in user-friendly real-time queriable platforms, these initiatives identified pathogenic variants in small nuclear RNA (snRNA) genes in 46 cases, a remarkable 2.69% of cases. These findings have allowed the translation in a relatively short time directly into clinical care, including reproductive counseling and personalized treatment strategies. Diagnoses were achieved in more than 25 hospitals across 13 of Spain’s 17 autonomous communities, milestones unlikely to have been reached independently. Notably, the rapid identification of conditions such as ReNU and other related syndromes underscores the value of URD Programs and international data-sharing frameworks to accelerate translational diagnostics.

Altogether, this work illustrates the diagnostic blind spots of exome sequencing and highlights the need for genome-wide approaches that capture noncoding and regulatory variation. All pathogenic snRNA variants uncovered here reside in noncoding regions of remarkable functional importance, highly conserved and depleted of variation. The diagnostic outcomes achieved were only possible through national-scale coordination, rapid data exchange, and real-time preprint communication. Looking ahead, integrating methylation, splicing, and structural modeling will likely unveil additional mechanisms underlying noncoding pathogenicity, not only in NDDs but potentially across a broader spectrum of Mendelian and complex diseases.

## Data Availability

All data produced in the present work are contained in the manuscript. Clinical and genomic data from patients are available to all participants in the Spanish URD Programs through specific databases.

## Acknowledgements

We thank all participants and families involved in the Spanish Undiagnosed Rare Disease Programs: “Infraestructura de Medicina de Precisión asociada a la Ciencia y la Tecnología en Medicina Genómica (IMPaCT-Genómica)”, “Programes de Malalties Rares no Diagnosticades de Catalunya (URD-Cat)” and “Programa de Enfermedades No Diagnosticadas (ENoD-CIBERER)”. IMPaCT-Genómica was supported by Instituto de Salud Carlos III, Ministerio de Ciencia e Innovación and the European Union European Regional Development Fund (IMP/00009), URD-Cat was supported by the Department of Health of Catalonia (Grant SLT002/16/00174). ENoD-CIBERER Program was funded by the Biomedical Network Research Center for Rare Diseases-CIBER-ER-ISCIII.

## Statement on Patient Identifiability

All potentially identifying information related to patients has been removed from this version manuscript and supplementary materials. This includes the removal of pedigrees, facial photographs, and any personal or clinical data that could lead to the identification of individuals (e.g., specific ages, family relationships, or clinical histories). Patient and sample identifiers used (e.g., U4-p1) are internal codes known only to members of the research group and are not recognizable by patients, families, or external collaborators. All case descriptions have been anonymized or removed to ensure full compliance with medRxiv’s patient privacy policy.

## References

1. Luque, J., Mendes, I., Gómez, B., Morte, B., López de Heredia, M., Herreras, E., Corrochano, V., Bueren, J., Gallano, P., Artuch, R., et al. (2022). CIBERER: Spanish national network for research on rare diseases: A highly productive collaborative initiative. Clin Genet 101, 481–493. 10.1111/cge.14113.

2. Bullich, G., Matalonga, L., Pujadas, M., Papakonstantinou, A., Piscia, D., Tonda, R., Artuch, R., Gallano, P., Garrabou, G., González, J.R., et al. (2022). Systematic Collaborative Reanalysis of Genomic Data Improves Diagnostic Yield in Neurologic Rare Diseases. The Journal of Molecular Diagnostics 24, 529–542. 10.1016/j.jmoldx.2022.02.003.

3. Gillentine, M.A., Wang, T., and Eichler, E.E. (2022). Estimating the Prevalence of De Novo Monogenic Neurodevelopmental Disorders from Large Cohort Studies. Biomedicines 10, 2865. 10.3390/biomedicines10112865.

4. Greene, D., Thys, C., Berry, I.R., Jarvis, J., Ortibus, E., Mumford, A.D., Freson, K., and Turro, E. (2024). Mutations in the U4 snRNA gene RNU4-2 cause one of the most prevalent monogenic neurodevelopmental disorders. Nat Med 30, 2165–2169. 10.1038/s41591-024-03085-5.

5. Greene, D., De Wispelaere, K., Lees, J., Codina-Solà, M., Jensson, B.O., Hales, E., Katrinecz, A., Nieto Molina, E., Pascoal, S., Pfundt, R., et al. (2025). Mutations in the small nuclear RNA gene RNU2-2 cause a severe neurodevelopmental disorder with prominent epilepsy. Nat Genet 57, 1367– 1373. 10.1038/s41588-025-02159-5.

6. Chen, Y., Dawes, R., Kim, H.C., Ljungdahl, A., Stenton, S.L., Walker, S., Lord, J., Lemire, G., Martin-Geary, A.C., Ganesh, V.S., et al. (2024). De novo variants in the RNU4-2 snRNA cause a frequent neurodevelopmental syndrome. Nature 632, 832–840. 10.1038/s41586-024-07773-7.

7. Nava, C., Cogne, B., Santini, A., Leitão, E., Lecoquierre, F., Chen, Y., Stenton, S.L., Besnard, T., Heide, S., Baer, S., et al. (2025). Dominant variants in major spliceosome U4 and U5 small nuclear RNA genes cause neurodevelopmental disorders through splicing disruption. Nat Genet 57, 1374–1388. 10.1038/s41588-025-02184-4.

8. Jackson, A., Thaker, N., Blakes, A., Rice, G., Griffiths-Jones, S., Balasubramanian, M., Campbell, J., Shannon, N., Choi, J., Hong, J., et al. (2025). Analysis of R-loop forming regions identifies RNU2-2 and RNU5B-1 as neurodevelopmental disorder genes. Nat Genet 57, 1362–1366. 10.1038/s41588-025-02209-y.

9. Li, D., Wang, Q., Bayat, A., Battig, M.R., Zhou, Y., Bosch, D.G.M., van Haaften, G., Granger, L., Petersen, A.K., Pérez-Jurado, L.A., et al. (2024). Spliceosome malfunction causes neurodevelopmental disorders with overlapping features. J Clin Invest 134. 10.1172/JCI171235.

10. Valenzuela, I., Codina-Solà, M., Vazquez, E., Cueto-González, A., Leno-Colorado, J., Lasa-Aranzasti, A., Trujillano, L., Masotto, B., Masas, M., Escobar, M., et al. (2024). Deep phenotyping of 11 individuals with pathogenic variants in RNU4-2 reveals a clinically recognizable syndrome. Genet Med 26, 101288. 10.1016/j.gim.2024.101288.

11. Schot, R., Ferraro, F., Geeven, G., Diderich, K.E.M., and Barakat, T.S. (2024). Re-analysis of whole genome sequencing ends a diagnostic odyssey: Case report of an RNU4-2 related neurodevelopmental disorder. Clin Genet 106, 512–517. 10.1111/cge.14574.

12. Mabin, J.W., Lewis, P.W., Brow, D.A., and Dvinge, H. (2021). Human spliceosomal snRNA sequence variants generate variant spliceosomes. RNA 27, 1186–1203. 10.1261/rna.078768.121.

13. Bousquets-Muñoz, P., Díaz-Navarro, A., Nadeu, F., Sánchez-Pitiot, A., López-Tamargo, S., Shuai, S., Balbín, M., Tubio, J.M.C., Beà, S., Martin-Subero, J.I., et al. (2022). PanCancer analysis of somatic mutations in repetitive regions reveals recurrent mutations in snRNA U2. NPJ Genom Med 7, 19. 10.1038/s41525-022-00292-2.

14. Newman, A.J., and Norman, C. (1992). U5 snRNA interacts with exon sequences at 5’ and 3’ splice sites. Cell 68, 743–754. 10.1016/0092-8674(92)90149-7.

15. Sontheimer, E.J., and Steitz, J.A. (1993). The U5 and U6 small nuclear RNAs as active site components of the spliceosome. Science 262, 1989–1996. 10.1126/science.8266094.

16. De Jonghe, J., Kim, H.C., Adedeji, A., Leitão, E., Dawes, R., Chen, Y., Blakes, A.J., Simons, C., Rius, R., Alvi, J.R., et al. (2025). Saturation genome editing of RNU4-2 reveals distinct dominant and recessive neurodevelopmental disorders. medRxiv, 2025.04.08.25325442. 10.1101/2025.04.08.25325442.

17. Rius, R., Blakes, A.J.M., Chen, Y., De Jonghe, J., Alvi, J.R., Amblard, F., Austin-Tse, C., Baer, S., Balasubramanian, M., Balton, E. V, et al. (2025). Biallelic variants in the non-coding RNA gene <em>RNU4-2</em> cause a recessive neurodevelopmental syndrome with distinct white matter changes. medRxiv, 2025.08.13.25333306. 10.1101/2025.08.13.25333306.

18. Gargano, M.A., Matentzoglu, N., Coleman, B., Addo-Lartey, E.B., Anagnostopoulos, A. V, Anderton, J., Avillach, P., Bagley, A.M., Bakštein, E., Balhoff, J.P., et al. (2024). The Human Phenotype Ontology in 2024: phenotypes around the world. Nucleic Acids Res 52, D1333–D1346. 10.1093/nar/gkad1005.

19. Laurie, S., Piscia, D., Matalonga, L., Corvó, A., Fernández-Callejo, M., Garcia-Linares, C., Hernandez-Ferrer, C., Luengo, C., Martínez, I., Papakonstantinou, A., et al. (2022). The RD-Connect Genome-Phenome Analysis Platform: Accelerating diagnosis, research, and gene discovery for rare diseases. Hum Mutat 43, 717–733. 10.1002/humu.24353.

20. Li, H. (2013). Aligning sequence reads, clone sequences and assembly contigs with BWA-MEM.

21. Vasimuddin, M., Misra, S., Li, H., and Aluru, S. (2019). Efficient Architecture-Aware Acceleration of BWA-MEM for Multicore Systems. In 2019 IEEE International Parallel and Distributed Processing Symposium (IPDPS), pp. 314–324. 10.1109/IPDPS.2019.00041.

22. Poplin, R., Ruano-Rubio, V., DePristo, M.A., Fennell, T.J., Carneiro, M.O., Van der Auwera, G.A., Kling, D.E., Gauthier, L.D., Levy-Moonshine, A., Roazen, D., et al. (2018). Scaling accurate genetic variant discovery to tens of thousands of samples. bioRxiv, 201178. 10.1101/201178.

23. Chen, X., Schulz-Trieglaff, O., Shaw, R., Barnes, B., Schlesinger, F., Källberg, M., Cox, A.J., Kruglyak, S., and Saunders, C.T. (2016). Manta: rapid detection of structural variants and indels for germline and cancer sequencing applications. Bioinformatics 32, 1220–1222. 10.1093/bioinformatics/btv710.

24. Boeva, V., Popova, T., Bleakley, K., Chiche, P., Cappo, J., Schleiermacher, G., Janoueix-Lerosey, I., Delattre, O., and Barillot, E. (2012). Control-FREEC: a tool for assessing copy number and allelic content using next-generation sequencing data. Bioinformatics 28, 423–425. 10.1093/bioinformatics/btr670.

25. Cameron, D.L., Schröder, J., Penington, J.S., Do, H., Molania, R., Dobrovic, A., Speed, T.P., and Papenfuss, A.T. (2017). GRIDSS: sensitive and specific genomic rearrangement detection using positional de Bruijn graph assembly. Genome Res 27, 2050–2060. 10.1101/GR.222109.117.

26. Dolzhenko, E., Deshpande, V., Schlesinger, F., Krusche, P., Petrovski, R., Chen, S., Emig-Agius, D., Gross, A., Narzisi, G., Bowman, B., et al. (2019). ExpansionHunter: a sequence-graph-based tool to analyze variation in short tandem repeat regions. Bioinformatics 35, 4754–4756. 10.1093/bioinformatics/btz431.

27. Robinson, J.T., Thorvaldsdóttir, H., Winckler, W., Guttman, M., Lander, E.S., Getz, G., and Mesirov, J.P. (2011). Integrative genomics viewer. Nat Biotechnol 29, 24–26. 10.1038/nbt.1754.

28. Lorenz, R., Bernhart, S.H., Höner Zu Siederdissen, C., Tafer, H., Flamm, C., Stadler, P.F., and Hofacker, I.L. (2011). ViennaRNA Package 2.0. Algorithms Mol Biol 6, 26. 10.1186/1748-7188-6-26.

29. Turunen, J.J., Niemelä, E.H., Verma, B., and Frilander, M.J. (2013). The significant other: splicing by the minor spliceosome. Wiley Interdiscip Rev RNA 4, 61–76. 10.1002/wrna.1141.

30. Gurovich, Y., Hanani, Y., Bar, O., Nadav, G., Fleischer, N., Gelbman, D., Basel-Salmon, L., Krawitz, P.M., Kamphausen, S.B., Zenker, M., et al. (2019). Identifying facial phenotypes of genetic disorders using deep learning. Nat Med 25, 60–64. 10.1038/s41591-018-0279-0.

31. Hsieh, T.-C., Bar-Haim, A., Moosa, S., Ehmke, N., Gripp, K.W., Pantel, J.T., Danyel, M., Mensah, M.A., Horn, D., Rosnev, S., et al. (2022). GestaltMatcher facilitates rare disease matching using facial phenotype descriptors. Nat Genet 54, 349–357. 10.1038/s41588-021-01010-x.

32. Min, J.L., Hemani, G., Davey Smith, G., Relton, C., and Suderman, M. (2018). Meffil: efficient normalization and analysis of very large DNA methylation datasets. Bioinformatics 34, 3983–3989. 10.1093/bioinformatics/bty476.

33. Aryee, M.J., Jaffe, A.E., Corrada-Bravo, H., Ladd-Acosta, C., Feinberg, A.P., Hansen, K.D., and Irizarry, R.A. (2014). Minfi: a flexible and comprehensive Bioconductor package for the analysis of Infinium DNA methylation microarrays. Bioinformatics 30, 1363–1369. 10.1093/bioinformatics/btu049.

34. Fortin, J.-P., Labbe, A., Lemire, M., Zanke, B.W., Hudson, T.J., Fertig, E.J., Greenwood, C.M.T., and Hansen, K.D. (2014). Functional normalization of 450k methylation array data improves replication in large cancer studies. Genome Biol 15, 503. 10.1186/s13059-014-0503-2.

35. Leek, J.T., Johnson, W.E., Parker, H.S., Jaffe, A.E., and Storey, J.D. (2012). The sva package for removing batch effects and other unwanted variation in high-throughput experiments. Bioinformatics 28, 882–883. 10.1093/bioinformatics/bts034.

36. Ho, D., Imai, K., King, G., Stuart, E., and Greifer, N. (2005). MatchIt: Nonparametric Preprocessing for Parametric Causal Inference. CRAN: Contributed Packages. 10.32614/CRAN.PACKAGE.MATCHIT.

37. Pelegí-Sisó, D., de Prado, P., Ronkainen, J., Bustamante, M., and González, J.R. (2021). methylclock: a Bioconductor package to estimate DNA methylation age. Bioinformatics 37, 1759–1760. 10.1093/bioinformatics/btaa825.

38. Ritchie, M.E., Phipson, B., Wu, D., Hu, Y., Law, C.W., Shi, W., and Smyth, G.K. (2015). limma powers differential expression analyses for RNA-sequencing and microarray studies. Nucleic Acids Res 43, e47–e47. 10.1093/NAR/GKV007.

39. Vos, N., Haghshenas, S., van der Laan, L., Russel, P.K.M., Rooney, K., Levy, M.A., Relator, R., Kerkhof, J., McConkey, H., Maas, S.M., et al. (2024). The detection of a strong episignature for Chung-Jansen syndrome, partially overlapping with Börjeson-Forssman-Lehmann and White-Kernohan syndromes. Hum Genet 143, 761–773. 10.1007/s00439-024-02679-w.

40. Meyer, D., Dimitriadou, E., Hornik, K., Weingessel, A., and Leisch, F. (2024). Misc Functions of the Department of Statistics, Probability Theory Group (Formerly: E1071), TU Wien [R package e1071 version 1.7-16]. CRAN: Contributed Packages. 10.32614/CRAN.PACKAGE.E1071.

41. Peters, T.J., Buckley, M.J., Statham, A.L., Pidsley, R., Samaras, K., V Lord, R., Clark, S.J., and Molloy, P.L. (2015). De novo identification of differentially methylated regions in the human genome. Epigenetics Chromatin 8, 1–16. 10.1186/1756-8935-8-6/TABLES/1.

42. Phipson, B., Maksimovic, J., and Oshlack, A. (2016). missMethyl: an R package for analyzing data from Illumina’s HumanMethylation450 platform. Bioinformatics 32, 286–288. 10.1093/bioinformatics/btv560.

43. Reichert, S.C., Li, R., A. Turner, S., van Jaarsveld, R.H., Massink, M.P.G., van den Boogaard, M.-J.H., del Toro, M., Rodríguez-Palmero, A., Fourcade, S., Schlüter, A., et al. (2020). HNRNPH1-related syndromic intellectual disability: Seven additional cases suggestive of a distinct syndromic neurodevelopmental syndrome. Clin Genet 98, 91–98. 10.1111/cge.13765.

44. Quinodoz, M., Rodenburg, K., Cvackova, Z., Kaminska, K., de Bruijn, S.E., Iglesias-Romero, A.B., Boonen, E.G.M., Ullah, M., Zomer, N., Folcher, M., et al. (2025). De novo and inherited dominant variants in U4 and U6 snRNAs cause retinitis pigmentosa. medRxiv, 2025.01.06.24317169. 10.1101/2025.01.06.24317169.

45. Cornelia de Lange Syndrome - GeneReviews® - NCBI Bookshelf https://www.ncbi.nlm.nih.gov/books/NBK1104/.

46. Veitia, R.A., Caburet, S., and Birchler, J.A. (2018). Mechanisms of Mendelian dominance. Clin Genet 93, 419–428. 10.1111/CGE.13107.

47. Putoux, A., Alqahtani, A., Pinson, L., Paulussen, A.D.C., Michel, J., Besson, A., Mazoyer, S., Borg, I., Nampoothiri, S., Vasiljevic, A., et al. (2016). Refining the phenotypical and mutational spectrum of Taybi-Linder syndrome. Clin Genet 90, 550–555. 10.1111/cge.12781.

48. Goumy, C., Gay-Bellile, M., Salaun, G., Kemeny, S., Eymard-Pierre, E., Biard, M., Pebrel-Richard, C., Vanlieferinghen, P., Francannet, C., Tchirkov, A., et al. (2016). A novel 2q14.1q14.3 deletion involving GLI2 and RNU4ATAC genes associated with partial corpus callosum agenesis and severe intrauterine growth retardation. Birth Defects Res A Clin Mol Teratol 106, 793–797. 10.1002/bdra.23535.

49. Chen, Y.-S., He, J.-F., Quan, T., Li, S.-B., and Li, D.-Z. (2023). Hydrocephalus and Growth Retardation: A Fetal RNU4ATAC-opathy Missed by Whole-Exome Sequencing. Mol Syndromol 13, 522–526. 10.1159/000524501.

50. Kim, J.-S., He, X., Liu, J., Duan, Z., Kim, T., Gerard, J., Kim, B., Pillai, M.M., Lane, W.S., Noble, W.S., et al. (2019). Systematic proteomics of endogenous human cohesin reveals an interaction with diverse splicing factors and RNA-binding proteins required for mitotic progression. Journal of Biological Chemistry 294, 8760–8772. 10.1074/jbc.RA119.007832.

51. Singh, A.K., Chen, Q., Nguyen, C., Meerzaman, D., and Singer, D.S. (2025). Cohesin regulates alternative splicing. Sci Adv 9, eade3876. 10.1126/sciadv.ade3876.

52. Boesler, C., Rigo, N., Anokhina, M.M., Tauchert, M.J., Agafonov, D.E., Kastner, B., Urlaub, H., Ficner, R., Will, C.L., and Lührmann, R. (2016). A spliceosome intermediate with loosely associated tri-snRNP accumulates in the absence of Prp28 ATPase activity. Nat Commun 7, 11997. 10.1038/ncomms11997.

