## Supplemental figures for "Characterization of snRNA-related neurodevelopmental disorders through the Spanish Undiagnosed Rare Disease Programs"

Sevilla-Porras et al.

**SUPPLEMENTAL TABLES**

**Table S1**: Primer sequences used for Sanger validation of *RNU4-2*, *RNU2-2*, and *RNU5B-1* variants.

**Table S2**: List of patients included in the Face2Gene analysis.

**Table S3**: List of patients' samples used for methylation analysis, including GSE accession numbers and matching parameters. Column Batch refers to patient designation as training or validation for the analysis.

**Table S4**: Metadata of patients with *de novo* variants in *RNU4-2*, *RNU2-2*, and *RNU5B-1* with structural domain annotation.

**Table S5**: Rare variants identified in *RNU4-2*, *RNU2-2*, and *RNU5B-1*, in our cohort with inheritance confirmation, gnomAD frequencies, variant classification, domain annotation and clinical classification.

**Table S6**: List of snRNA genes of the major spliceosome included in the analysis of biallelic variants. Genes were selected based on their expression levels in brain tissue, according to GTEx data.

**Table S7**: Detailed list of biallelic snRNA variants identified in cases and controls. Information on gnomAD frequencies, zygosity, and associated phenotype is included.

**Table S8**: Clinical features of patients with *de novo* snRNA variants, annotated with Human Phenotype Ontology (HPO), grouped by major HPO categories. These terms were used for reverse phenotyping and subsequently for clustering analysis. Numbers in parentheses represent total responses per HPO term.

**Table S9**: Detailed prenatal history of patients with variants in *RNU2-2*, *RNU4-2* and *RNU5B-1*. * One case conceived by egg donation resulting in twin pregnancy with ulterior single-twin intrauterine loss in the first trimester. ** Two male patients who are a monozygotic twin pair. *** A male patient large for gestational age (birth weight of 4,535 grams) was excluded due to maternal gestational diabetes. Prenatal diagnosis in the *RNU4-2* group included two cases with prenatal array CGH and one chorionic villus sampling performed due to increased nuchal translucency and ventriculomegaly. All results showed a normal karyotype (46,XX) and normal array CGH findings. Fetal mild ventriculomegaly was detected in five patients, and in one case, an MRI at 32 weeks also revealed a cavum septum pellucidum cyst and flattened temporal lobe morphology.

**Table ST0**: List of differentially methylated probes (DMPs) in *RNU4-2* patients.

**Table S11**: Summary of differentially methylated regions (DMRs), including genomic coordinates and overlapping genes.

### SUPPLEMENTAL FIGURES

**Figure S1: Pedigrees of patients with rare *de novo* variants in *RNU4-2* and *RNU5B-1*.** (Panel A) *RNU4-2* variants: A = n.64_65insT, B = n.66A>G, C = n.67A>G, D = n.69C>T, E = n.69C>G, F = n.77_78insC, G = n.77_78insG, H = n.78A>C. (Panel B) *RNU5B-1* variants: 1 = n.37G>C, 2 = n.39C>G, 3 = n.42_43insA. ***Removed for preprint versions***

**Figure S2: Bar plot showing the distribution of identified variants in the *RNU4-2* (Panel A) and *RNU2-2* (Panel B) genes within the IMPaCT-Genómica, ENoD-CIBERER, URD-Cat cohorts, filtered by population allele count <40 (gnomAD v4.1.0).** The legend denotes the inheritance pattern of the identified variants. The x-axis also indicates the number of unaffected individuals from the internal cohort carrying the variants (red) and the number of carriers in gnomAD v4 (blue).

**Figure S3: Bar plot showing the distribution of identified variants in *RNU5B-1* within the IMPaCT-Genómica, ENoD-CIBERER, URD-Cat cohort, filtered for a population allele count (gnomAD v4.1.0) below 40.** The legend denotes the inheritance status of the variants identified. The x-axis also indicates the number of unaffected individuals from the internal cohort carrying the variants (red) and the number of carriers in gnomAD v4 (blue).

**Figure S4: Expression heatmap of snRNA genes across human tissues based on GTEx data.** For each snRNA subfamily (RNU2, RNU4, RNU5, RNU6), the top 10 subunits with the highest expression in brain tissues are displayed, along with selected non-neural tissues (liver, pancreas, and whole blood) for comparison. The color scale represents relative expression levels, with blue indicating higher expression and yellow indicating lower expression.

**Figure S5: Visualization of digenic variant combinations observed in the cohort.** A. Each circle represents a pairwise combination of snRNA subunits. Circles are divided into two coloured segments: red for individuals with neurodevelopmental disorders (NDD) and blue for unaffected individuals. The number inside each segment indicates the number of individuals carrying the respective digenic combination in each group. B. Comparison of the overall number of individuals carrying digenic combinations between NDD and unaffected groups. A Fisher's exact test was applied to assess statistical significance ($p < 0.05$; indicated by *).

**Figure S6. Pedigree of family U4-p30 carrying compound heterozygous variants in *RNU4-2*.** The proband is indicated with an arrow. Individuals shaded in black are affected with a retinitis pigmentosa phenotype, and genotypes are shown below each tested individual. ***Removed for preprint versions***

**Figure S7: Predicted RNA secondary structures for wild-type vs. mutant *RNU5B-1* (panel A), *RNU2-2* (panel B) and RNU4-1 (panel C) and *RNU4-2* (panel D) transcripts using RNAfold.** Minimum free energy (MFE) and centroid structure drawings are shown along mountain plot representation of MFE, thermodynamic ensemble of RNA structures (pf) and centroid structure. Structure drawings are coloured by base-pairing probabilities, and mountain plot includes positional entropy for each nucleotide. Location of the variants is indicated by a blue arrow. Folding constraint of sm domain 9 positions not to be paired was specified to create the prediction.

**Figure S8: Heatmap from hierarchical clustering of phenotypic profiles based on HPO terms.** The horizontal axis represents individual patient cases, along with their corresponding ages and pathogenic variants in the genes *RNU2-2*, *RNU4-2*, and *RNU5B-1*. The vertical axis categorizes the HPO terms for each patient into four groups: facial dysmorphologies, musculoskeletal and growth abnormalities, nervous system physiology, and other systems. Clustering was performed using the Euclidean method, indicating the presence (red), absence (blue), or missing data (white) of HPO terms. The analysis reveals patterns of similarity, highlighting correlations between phenotype and genotype.

**Figure S9: Facial photographs of 14 individuals with pathogenic *de novo* variants in *RNU4-2* (panel A) and 4 individuals with pathogenic *de novo* variants in *RNU5B-1* (panel B).** The estimated age at the time each photograph was taken is indicated above each image. **** Removed for preprint versions ****

**Figure S10: Supplementary figures of *RNU4-2* episignature analysis.** A. MDS plots from the LOOCV approach for U4 episignature identification, with case, control, and test samples shown in red, blue, and yellow, respectively. B. Probability classification results from the constructed SVM model for the U4 episignature. Samples are categorized by genes, with colors indicating specific variants. C. Correlation plot comparing methylation differences (Δβ) for episignature-specific probes previously reported. Each point represents a CpG probe from the *RNU4-2* episignature described in the previous study. The x-axis shows the Δβ values from our current analysis, and the y-axis shows the Δβ values reported in the original publication. Colour indicates whether the direction of methylation change is concordant between studies. Pearson correlation coefficient is also calculated D. Heatmap showing histone mark enrichment analysis of the top 500 CpGs derived from the U4 episignature analysis. Enrichments were calculated using ChIP-seq data from 18 blood cell type-related epigenomes provided by the Roadmap Epigenomics Consortium. The color scale represents the odds ratio of significant CpG sites, as determined by one-sided Fisher's exact tests using filtered EPICv2 probes as background. E. Heatmap illustrating chromatin state enrichment for the same top 500 CpGs, based on annotations from the ChromHMM expanded 18-state model (NIH Roadmap Epigenomics). Color intensity corresponds to the odds ratio of significant enrichments across 18 chromatin states. F. Dot plot displaying the enrichment of transcription factor binding sites (TFBS) in the top 500 CpGs using data from the GTRD database (v18.06). Statistical significance was assessed using one-sided Fisher's exact tests with EPICv2 CpGs as background.

**Figure S11**. ***RNU2-2* and *RNU5B-1* episignature analysis** A. Hierarchical clustering using U2-specific probes. Rows correspond to selected probes, while columns represent samples. The color scale indicates methylation levels (blue: hypomethylated; red: hypermethylated). U2, U4, U5, and control groups are labeled in blue, pink, yellow, and gray, respectively. B. MDS plot from the LOOCV approach for U2 episignature identification. Samples are categorized into red (controls), blue (cases), and yellow (testing samples). C. Hierarchical clustering using U5-specific probes, following the same layout as panel A. D. MDS plot from the LOOCV approach for U5 episignature identification, with samples colored as in panel B.

**Figure S12. Differentially methylated regions (DMRs).** Upper panel: UCSC Genome Browser screenshot displaying the genomic context of the DMR, including annotated genes (blue), CpG islands (green), H3K27Ac marks, ENCODE cis regulatory elements (transcription start sites (TSS; red), enhancers (E; yellow), and weak enhancers (WE; pale yellow)) , GeneHancer annotations and JASPAR Transcription factor binding site. Lower panel: plot showing methylation levels (mean ± confidence intervals) for each CpG probe within the DMR in U4 cases (pink) and others (gray).

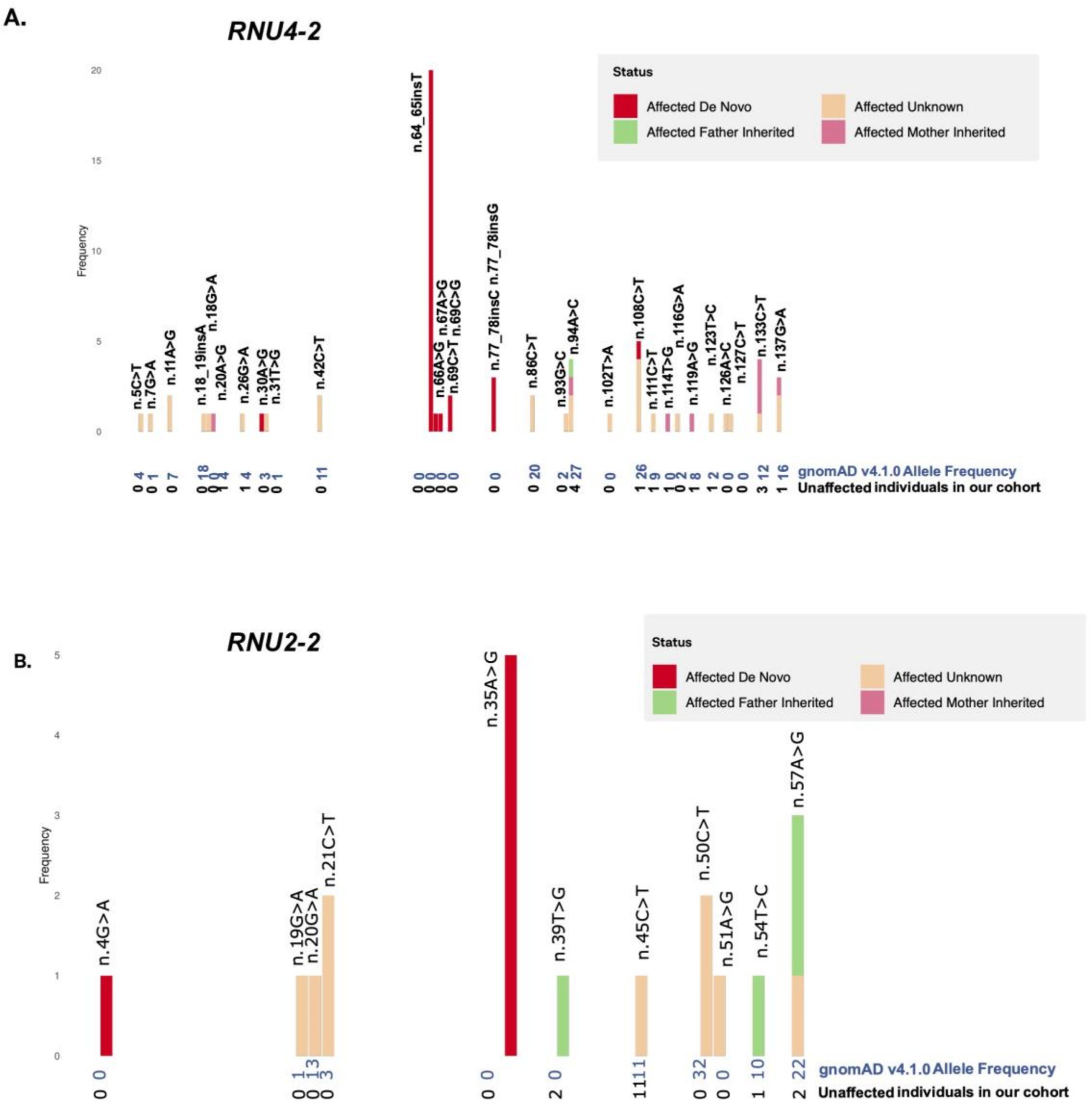
A.
RNU4-2
Status
Affected De Novo
Affected Unknown
Affected Father Inherited
Affected Mother Inherited
Frequency
n.5C>T
n.7G>A
n.11A>G
n.18_19insA
n.18G>A
n.20A>G
n.26G>A
n.30A>G
n.31T>G
n.42C>T
n.64_65insT
n.66A>G
n.67A>G
n.69C>T
n.69C>G
n.77_78insC
n.77_78insG
n.86C>T
n.93G>C
n.94A>C
n.102T>A
n.108C>T
n.111C>T
n.114T>G
n.116G>A
n.119A>G
n.123T>C
n.126A>C
n.127C>T
n.133C>T
n.137G>A
gnomAD v4.1.0 Allele Frequency
Unaffected individuals in our cohort
B.
RNU2-2
Status
Affected De Novo
Affected Unknown
Affected Father Inherited
Affected Mother Inherited
Frequency
n.4G>A
n.19G>A
n.20G>A
n.21C>T
n.35A>G
n.39T>G
n.45C>T
n.50C>T
n.51A>G
n.54T>C
n.57A>G
gnomAD v4.1.0 Allele Frequency
Unaffected individuals in our cohort

*Figure S 2*

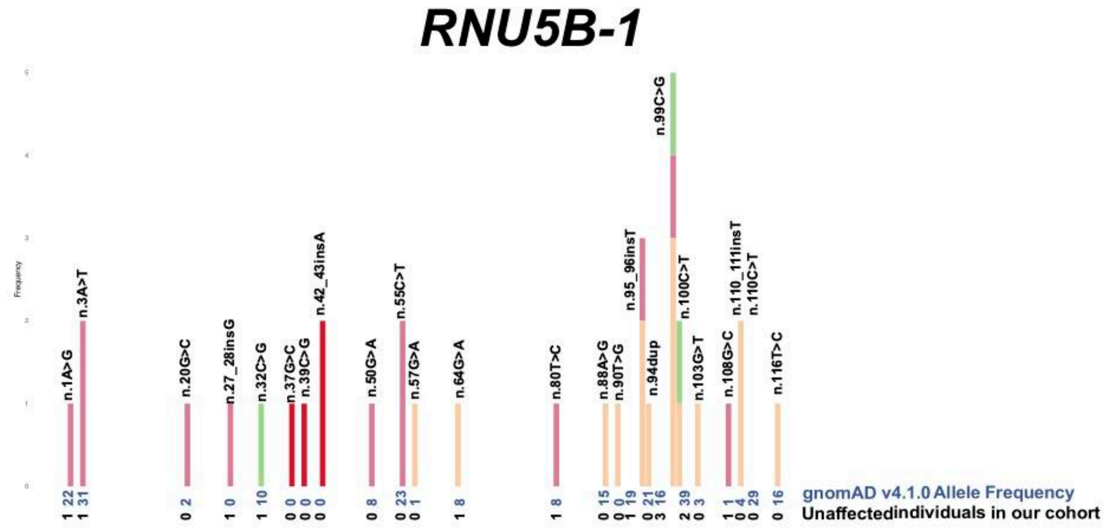


*Figure S 3*

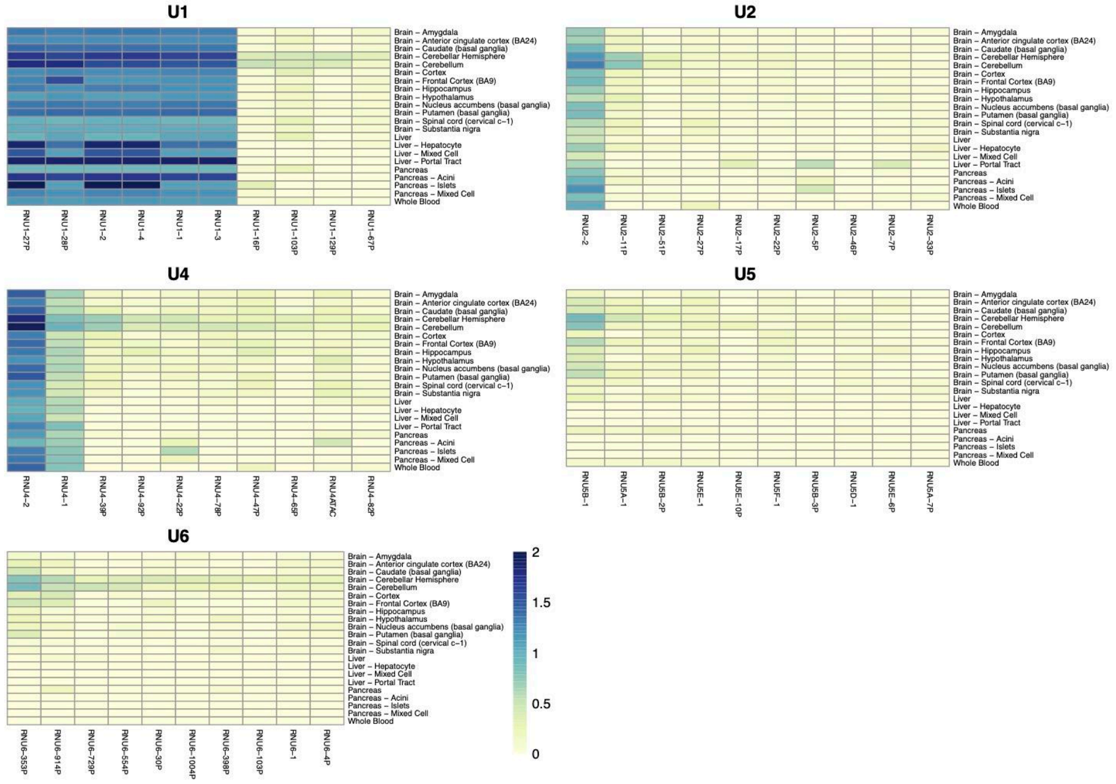


*Figure S 4*

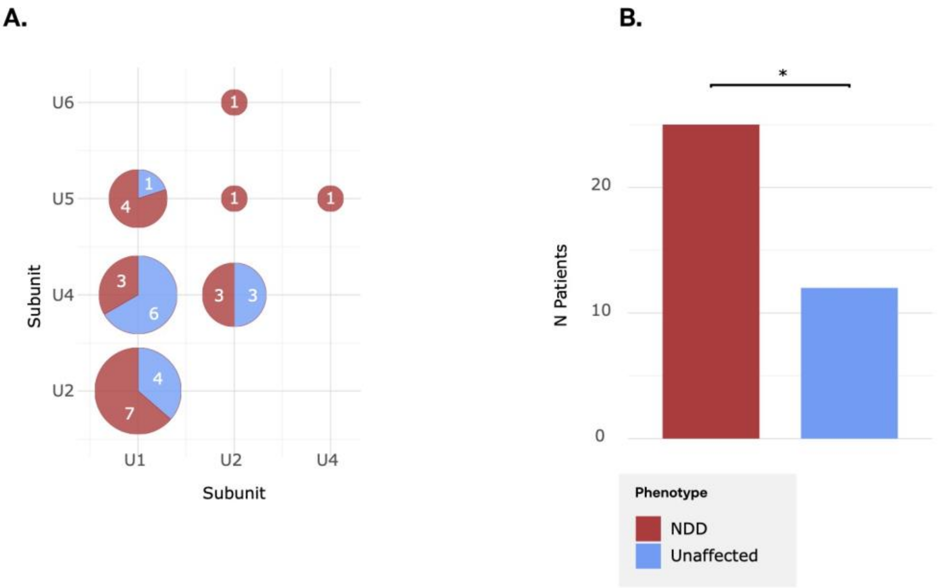
A.
U6
U5
U4
U2
Subunit
U1
U2
U4
Subunit
1
1
4
1
1
3
6
3
3
4
7
B.
*
N Patients
20
10
0
Phenotype
NDD
Unaffected

*Figure S 5*

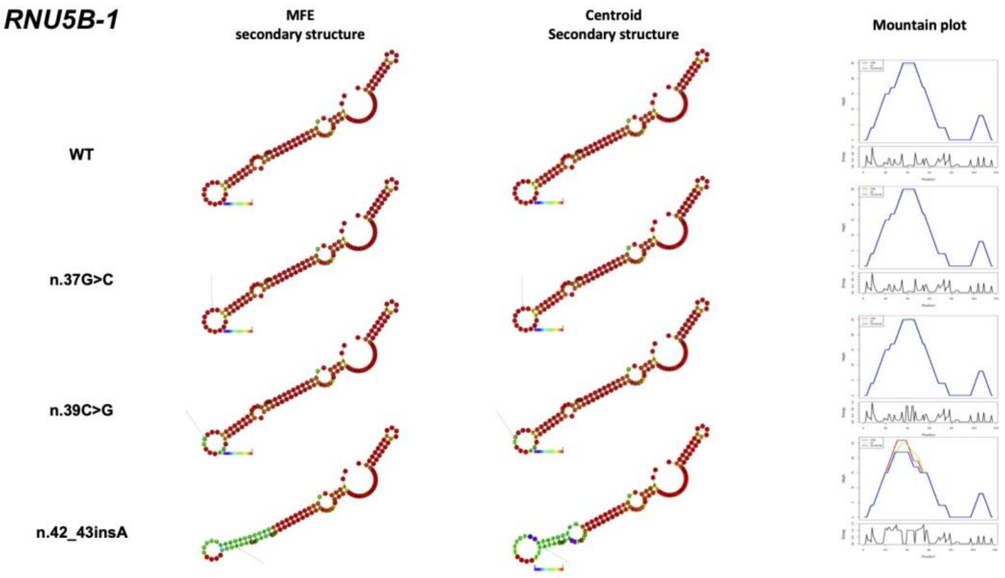
RNU5B-1
MFE
secondary structure
Centroid
Secondary structure
Mountain plot
WT
n.37G>C
n.39C>G
n.42_43insA

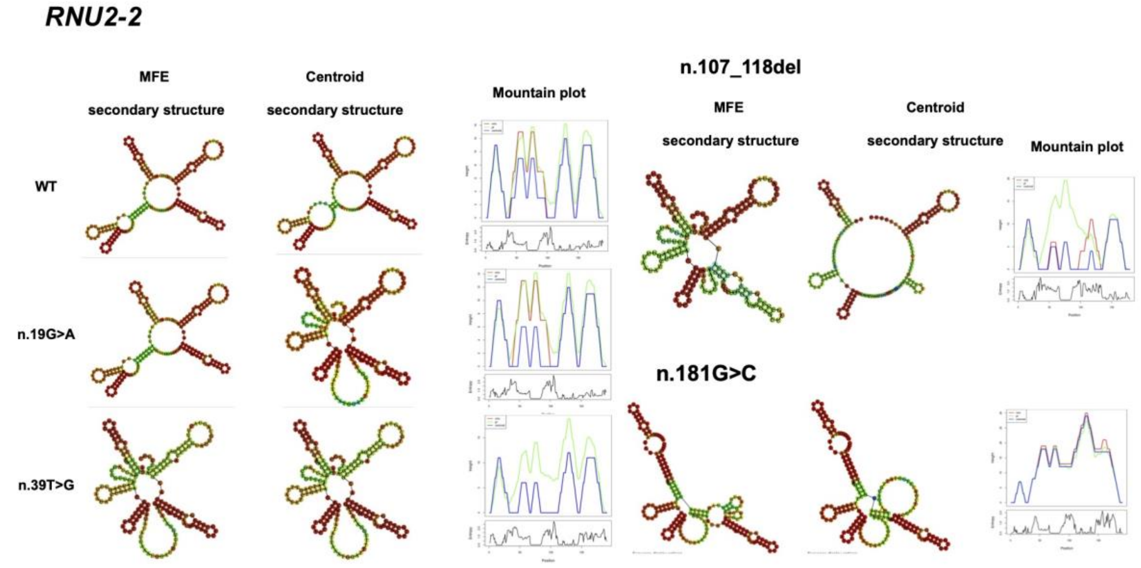
RNU2-2
MFE
secondary structure
Centroid
secondary structure
Mountain plot
WT
n.19G>A
n.39T>G
n.107_118del
MFE
secondary structure
Centroid
secondary structure
Mountain plot
n.181G>C

***RNU4-1***

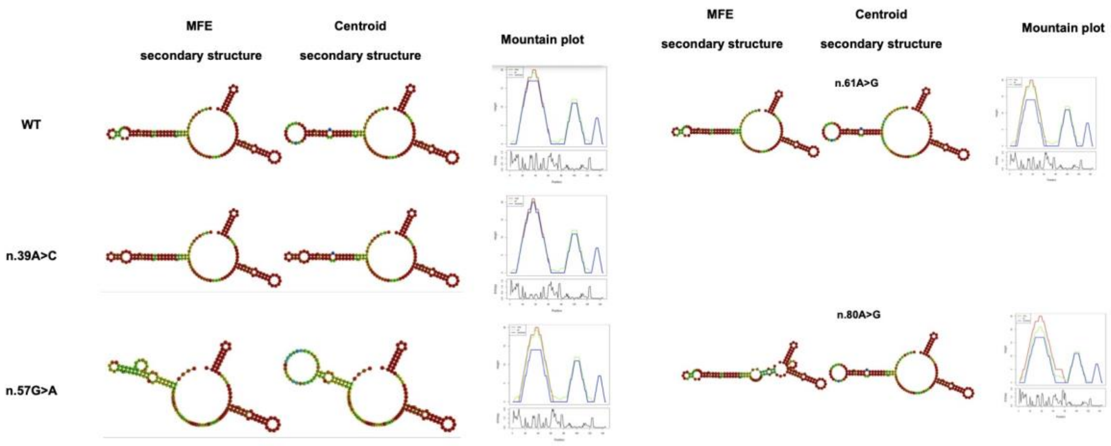


***RNU4-2***

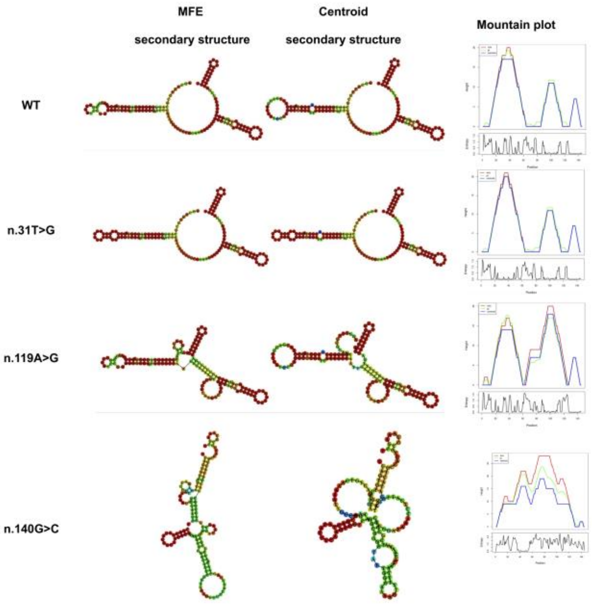


*Figure S 7*

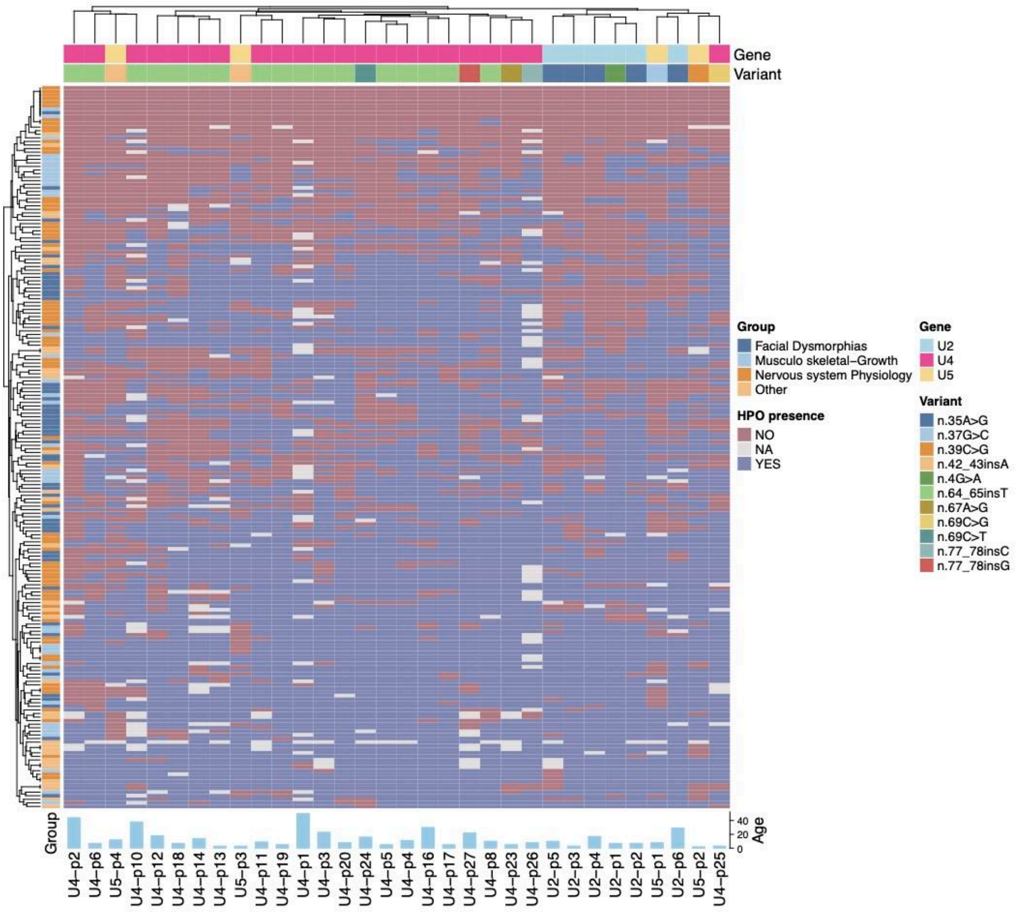
Gene
Variant
Group
Facial Dysmorphias
Musculo skeletal–Growth
Nervous system Physiology
Other
HPO presence
NO
NA
YES
Gene
U2
U4
U5
Variant
n.35A>G
n.37G>C
n.39C>G
n.42_43insA
n.4G>A
n.64_65insT
n.67A>G
n.69C>G
n.69C>T
n.77_78insC
n.77_78insG
Group
Age
40
20
0
U4–p2
U4–p6
U5–p4
U4–p10
U4–p12
U4–p18
U4–p14
U4–p13
U5–p3
U4–p11
U4–p19
U4–p1
U4–p3
U4–p20
U4–p24
U4–p5
U4–p4
U4–p16
U4–p17
U4–p27
U4–p8
U4–p23
U4–p26
U2–p5
U2–p3
U2–p4
U2–p1
U2–p2
U5–p1
U2–p6
U5–p2
U4–p25

*Figure S 8*

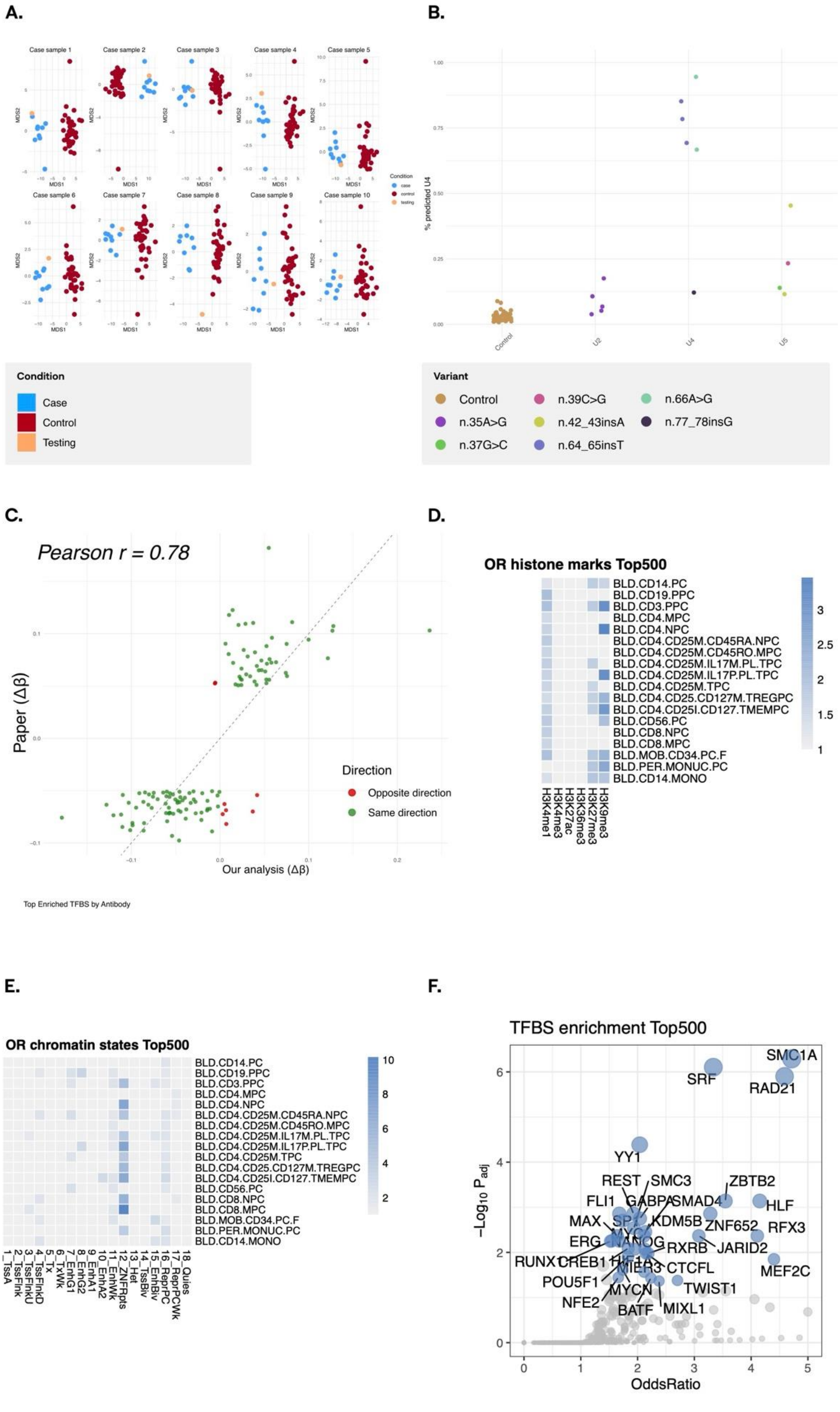
A.
Case sample 1
Case sample 2
Case sample 3
Case sample 4
Case sample 5
Case sample 6
Case sample 7
Case sample 8
Case sample 9
Case sample 10
MDS1
MDS2
Condition
Case
Control
Testing
B.
% predicted U4
Control
U2
U4
U5
Variant
Control
n.35A>G
n.37G>C
n.39C>G
n.42_43insA
n.64_65insT
n.66A>G
n.77_78insG
C.
Pearson r = 0.78
Paper (Δβ)
Our analysis (Δβ)
Direction
Opposite direction
Same direction
D.
OR histone marks Top500
BLD.CD14.PC
BLD.CD19.PPC
BLD.CD3.PPC
BLD.CD4.MPC
BLD.CD4.NPC
BLD.CD4.CD25M.CD45RA.NPC
BLD.CD4.CD25M.CD45RO.MPC
BLD.CD4.CD25M.IL17M.PL.TPC
BLD.CD4.CD25M.IL17P.PL.TPC
BLD.CD4.CD25M.TPC
BLD.CD4.CD25.CD127M.TREGPC
BLD.CD4.CD25I.CD127.TMEMPC
BLD.CD56.PC
BLD.CD8.NPC
BLD.CD8.MPC
BLD.MOB.CD34.PC.F
BLD.PER.MONUC.PC
BLD.CD14.MONO
H3K4me1
H3K4me3
H3K27ac
H3K36me3
H3K27me3
H3K9me3
Top Enriched TFBS by Antibody
E.
OR chromatin states Top500
1_TssA
2_TssFlnk
3_TssFlnkU
4_TssFlnkD
5_Tx
6_TxWk
7_EnhG1
8_EnhG2
9_EnhA1
10_EnhA2
11_EnhWk
12_ZNF/Rpts
13_Het
14_TssBiv
15_EnhBiv
16_ReprPC
17_ReprPCWk
18_Quies
F.
TFBS enrichment Top500
−Log10 Padj
OddsRatio
SMC1A
SRF
RAD21
YY1
REST
SMC3
ZBTB2
FLI1
GABPA
SMAD4
HLF
MAX
SPI1
KDM5B
ZNF652
RFX3
ERG
NANOG
RXRB
JARID2
RUNX1
CREB1
HIF1A
CTCFL
MEF2C
POU5F1
MYCN
TWIST1
NFE2
BATF
MIXL1

*Figure S 10*

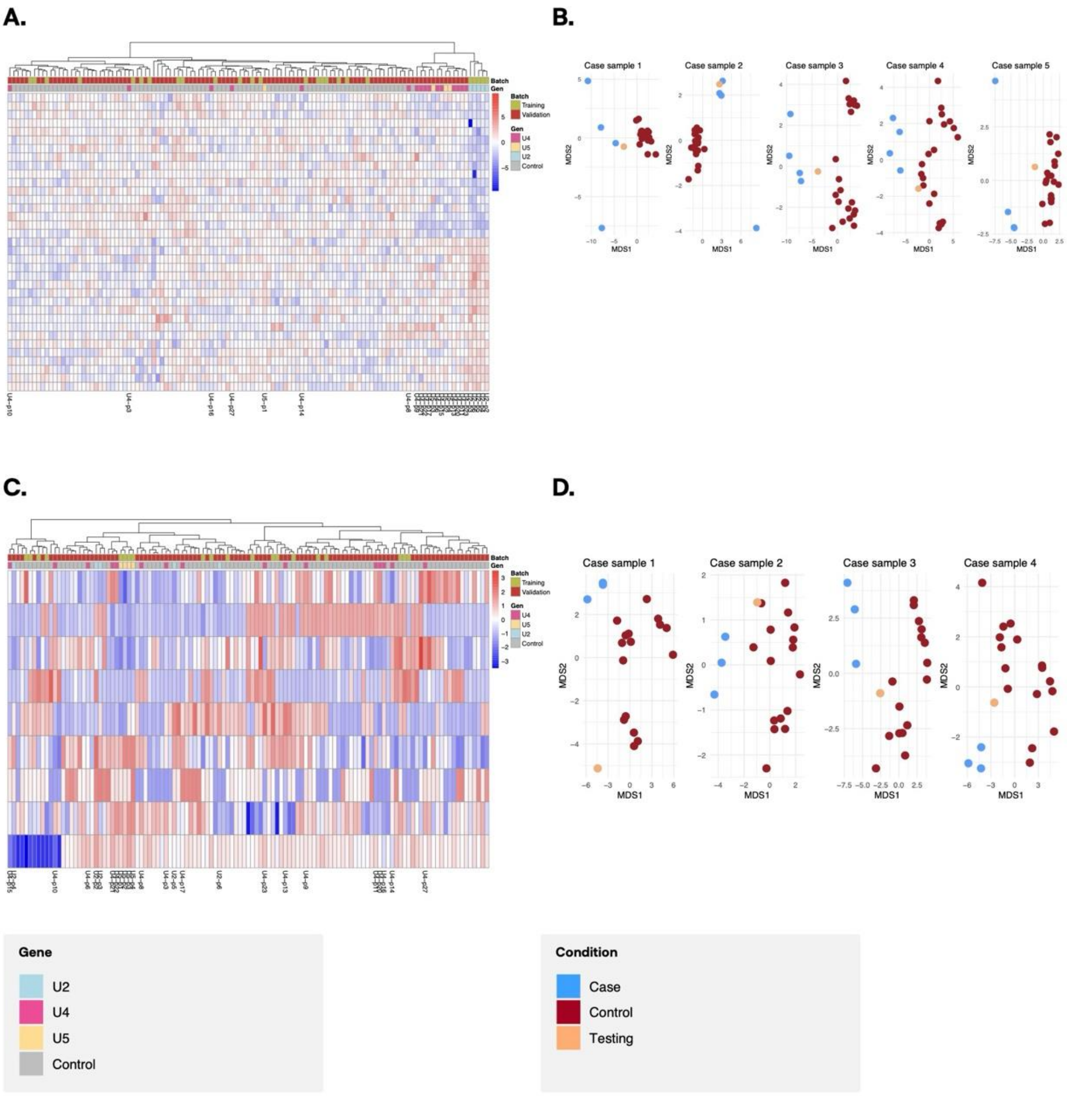
A.
B.
Case sample 1
Case sample 2
Case sample 3
Case sample 4
Case sample 5
MDS1
MDS2
Batch
Training
Validation
Gen
U4
U5
U2
Control
C.
D.
Case sample 1
Case sample 2
Case sample 3
Case sample 4
Gene
U2
U4
U5
Control
Condition
Case
Control
Testing

*Figure S 11*

A.

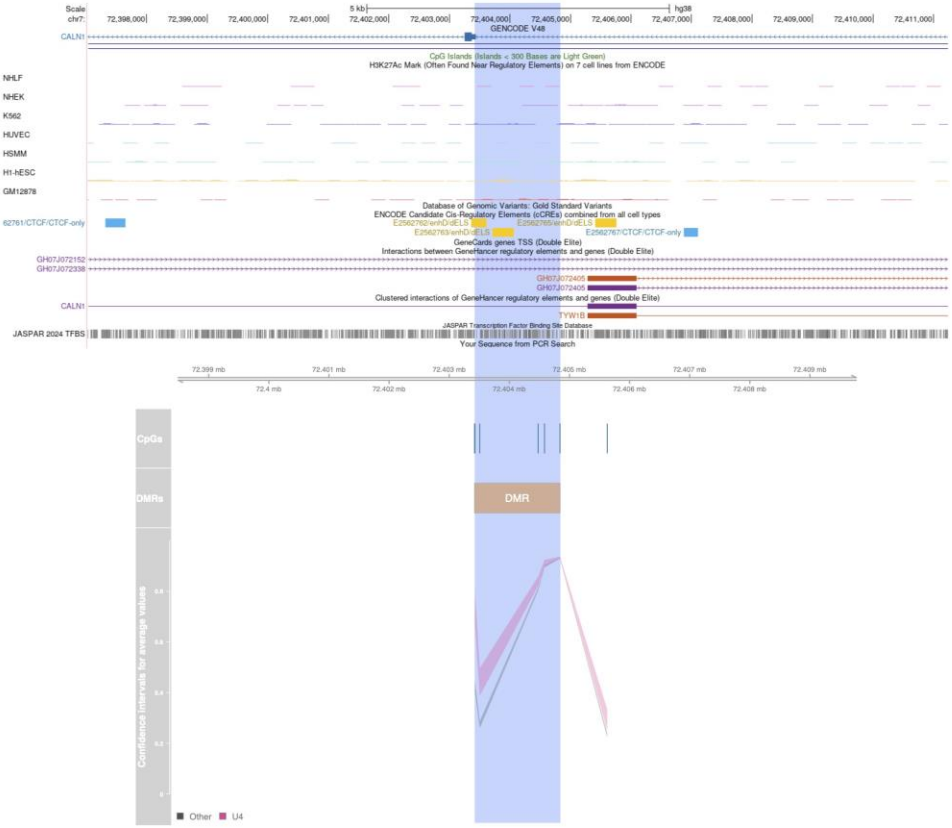
Scale
chr7:
5 kb
hg38
GENCODE V48
CALN1
CpG Islands (Islands < 300 Bases are Light Green)
H3K27Ac Mark (Often Found Near Regulatory Elements) on 7 cell lines from ENCODE
NHLF
NHEK
K562
HUVEC
HSMM
H1-hESC
GM12878
Database of Genomic Variants: Gold Standard Variants
ENCODE Candidate Cis-Regulatory Elements (cCREs) combined from all cell types
E2562761/CTCF/CTCF-only
E2562767/CTCF/CTCF-only
GeneCards genes TSS (Double Elite)
Interactions between GeneHancer regulatory elements and genes (Double Elite)
GH07J072152
GH07J072338
GH07J072405
Clustered interactions of GeneHancer regulatory elements and genes (Double Elite)
TYW1B
JASPAR 2024 TFBS
JASPAR Transcription Factor Binding Site Database
Your Sequence from PCR Search
CpGs
DMRs
DMR
Confidence intervals for average values
Other
U4

B.

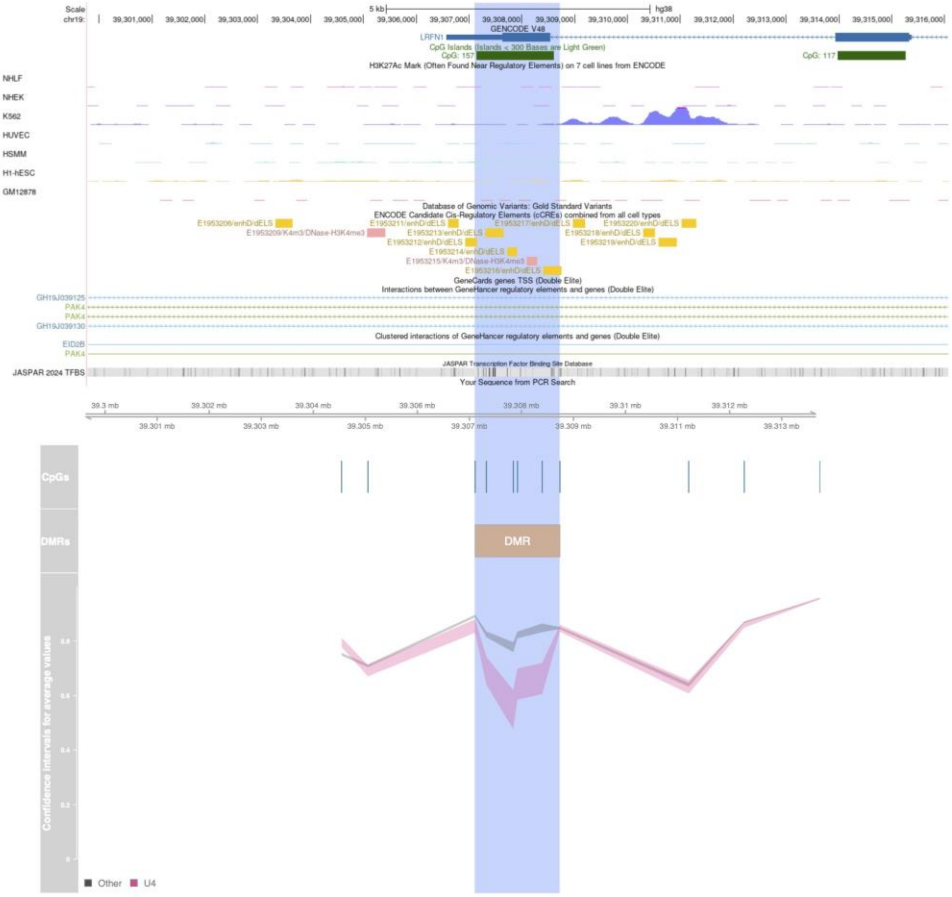
Scale
chr19:
5 kb
hg38
GENCODE V48
LRFN1
CpG Islands (Islands < 300 Bases are Light Green)
CpG: 157
CpG: 117
H3K27Ac Mark (Often Found Near Regulatory Elements) on 7 cell lines from ENCODE
NHLF
NHEK
K562
HUVEC
HSMM
H1-hESC
GM12878
Database of Genomic Variants: Gold Standard Variants
ENCODE Candidate Cis-Regulatory Elements (cCREs) combined from all cell types
GeneCards genes TSS (Double Elite)
Interactions between GeneHancer regulatory elements and genes (Double Elite)
GH19J039125
PAK4
GH19J039130
EID2B
Clustered interactions of GeneHancer regulatory elements and genes (Double Elite)
JASPAR 2024 TFBS
JASPAR Transcription Factor Binding Site Database
Your Sequence from PCR Search
CpGs
DMRs
DMR
Confidence intervals for average values
Other
U4

C.

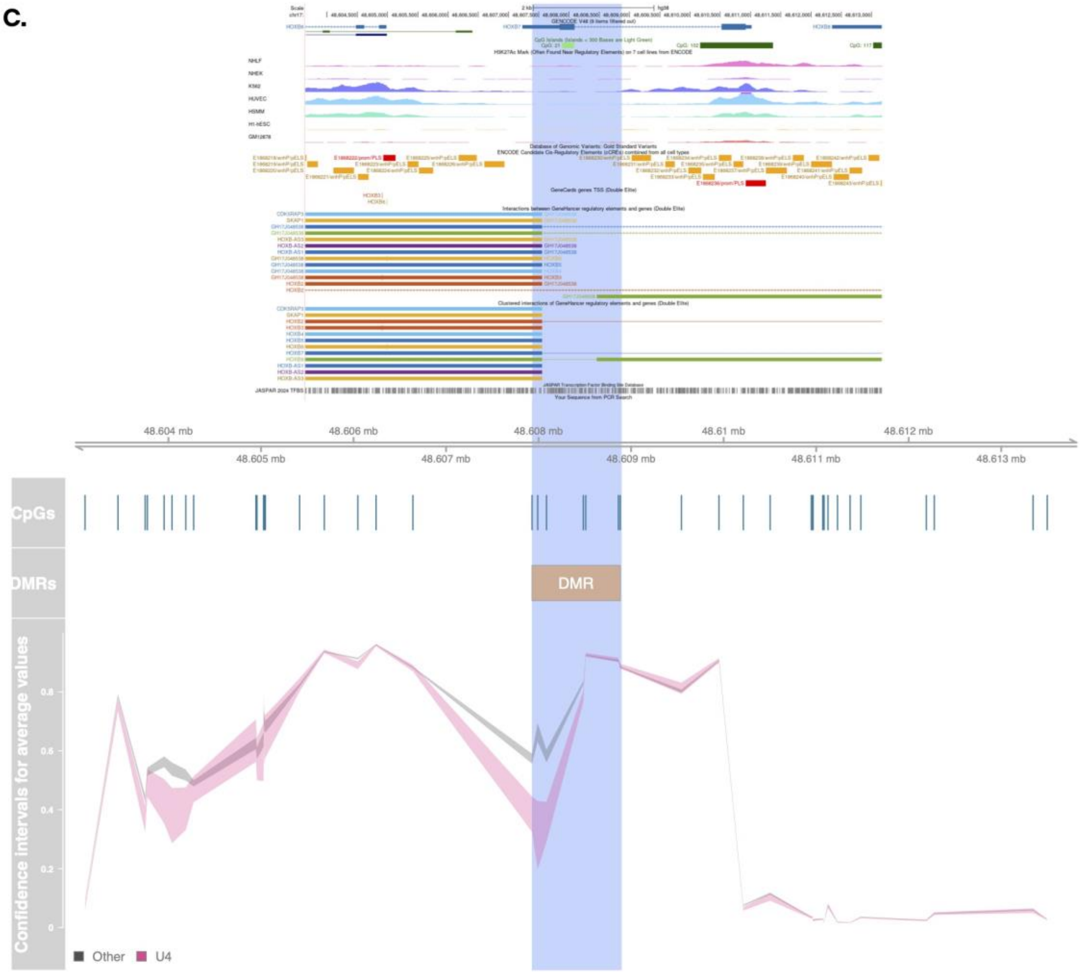

48.604 mb
48.605 mb
48.606 mb
48.607 mb
48.608 mb
48.609 mb
48.61 mb
48.611 mb
48.612 mb
48.613 mb
CpGs
DMRs
DMR
Confidence intervals for average values
0.8
0.6
0.4
0.2
0
Other
U4

D.

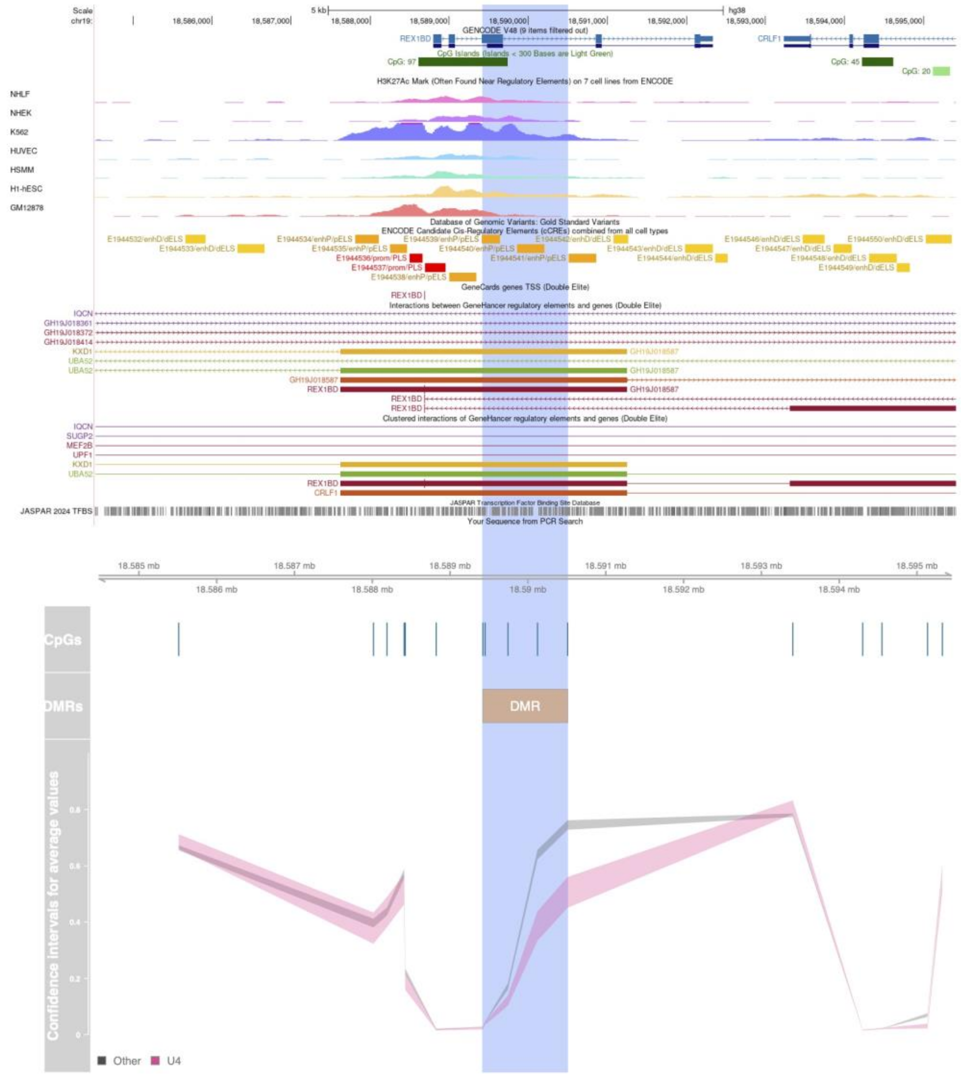

Scale
chr19:
5 kb
hg38
GENCODE V48 (9 items filtered out)
REX1BD
CRLF1
CpG Islands (Islands < 300 Bases are Light Green)
CpG: 97
CpG: 45
CpG: 20
H3K27Ac Mark (Often Found Near Regulatory Elements) on 7 cell lines from ENCODE
NHLF
NHEK
K562
HUVEC
HSMM
H1-hESC
GM12878
Database of Genomic Variants: Gold Standard Variants
ENCODE Candidate Cis-Regulatory Elements (cCREs) combined from all cell types
GeneCards genes TSS (Double Elite)
Interactions between GeneHancer regulatory elements and genes (Double Elite)
Clustered interactions of GeneHancer regulatory elements and genes (Double Elite)
JASPAR Transcription Factor Binding Site Database
JASPAR 2024 TFBS
Your Sequence from PCR Search
CpGs
DMRs
DMR
Confidence intervals for average values
Other
U4

**E.**

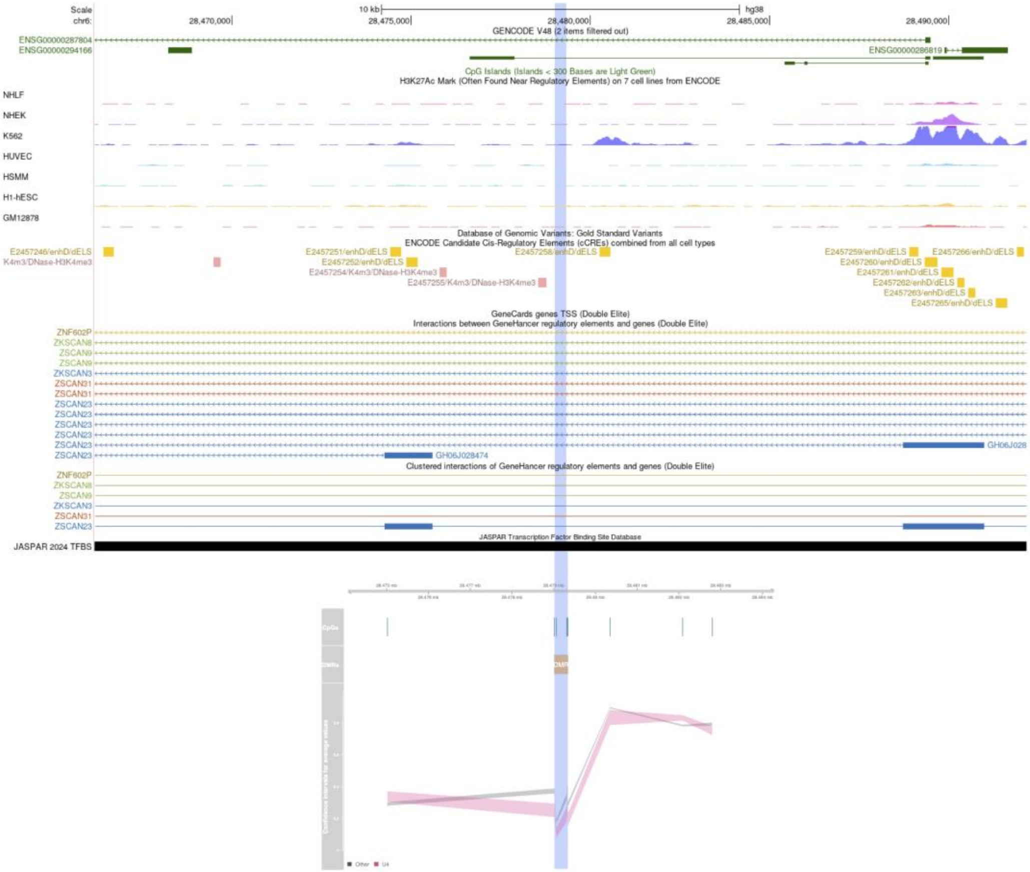
Scale
chr6:
10 kb
hg38
28,470,000
28,475,000
28,480,000
28,485,000
28,490,000
GENCODE V48 (2 items filtered out)
ENSG00000287804
ENSG00000294166
ENSG00000286819
CpG Islands (Islands < 300 Bases are Light Green)
H3K27Ac Mark (Often Found Near Regulatory Elements) on 7 cell lines from ENCODE
NHLF
NHEK
K562
HUVEC
HSMM
H1-hESC
GM12878
Database of Genomic Variants: Gold Standard Variants
ENCODE Candidate Cis-Regulatory Elements (cCREs) combined from all cell types
E2457246/enhD/dELS
K4m3/DNase-H3K4me3
E2457251/enhD/dELS
E2457252/enhD/dELS
E2457254/K4m3/DNase-H3K4me3
E2457255/K4m3/DNase-H3K4me3
E2457258/enhD/dELS
E2457259/enhD/dELS
E2457266/enhD/dELS
E2457260/enhD/dELS
E2457261/enhD/dELS
E2457262/enhD/dELS
E2457263/enhD/dELS
E2457265/enhD/dELS
GeneCards genes TSS (Double Elite)
Interactions between GeneHancer regulatory elements and genes (Double Elite)
ZNF602P
ZKSCAN8
ZSCAN9
ZSCAN9
ZKSCAN3
ZSCAN31
ZSCAN31
ZSCAN23
ZSCAN23
ZSCAN23
ZSCAN23
ZSCAN23
ZSCAN23
GH06J028474
GH06J028
Clustered interactions of GeneHancer regulatory elements and genes (Double Elite)
ZNF602P
ZKSCAN8
ZSCAN9
ZKSCAN3
ZSCAN31
ZSCAN23
JASPAR Transcription Factor Binding Site Database
JASPAR 2024 TFBS
CpGs
DMPs
DMP
Other
U4

**F.**

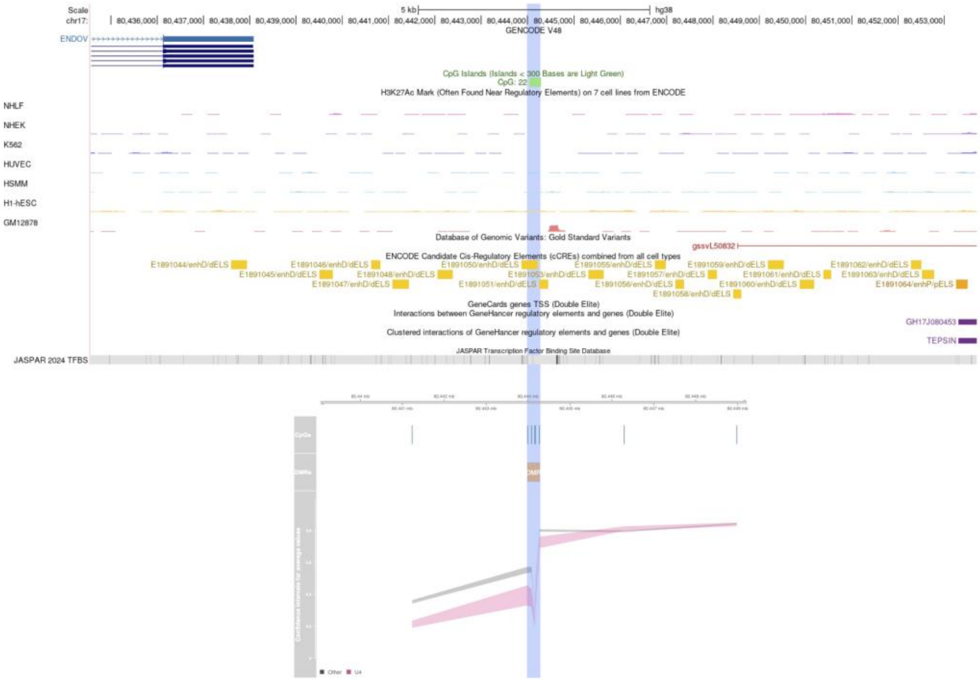
Scale
chr17:
5 kb
hg38
80,436,000
80,437,000
80,438,000
80,439,000
80,440,000
80,441,000
80,442,000
80,443,000
80,444,000
80,445,000
80,446,000
80,447,000
80,448,000
80,449,000
80,450,000
80,451,000
80,452,000
80,453,000
GENCODE V48
ENDOV
CpG Islands (Islands < 300 Bases are Light Green)
CpG: 22
H3K27Ac Mark (Often Found Near Regulatory Elements) on 7 cell lines from ENCODE
NHLF
NHEK
K562
HUVEC
HSMM
H1-hESC
GM12878
Database of Genomic Variants: Gold Standard Variants
gssvL50832
ENCODE Candidate Cis-Regulatory Elements (cCREs) combined from all cell types
GeneCards genes TSS (Double Elite)
Interactions between GeneHancer regulatory elements and genes (Double Elite)
GH17J080453
Clustered interactions of GeneHancer regulatory elements and genes (Double Elite)
TEPSIN
JASPAR Transcription Factor Binding Site Database
JASPAR 2024 TFBS

**G.**

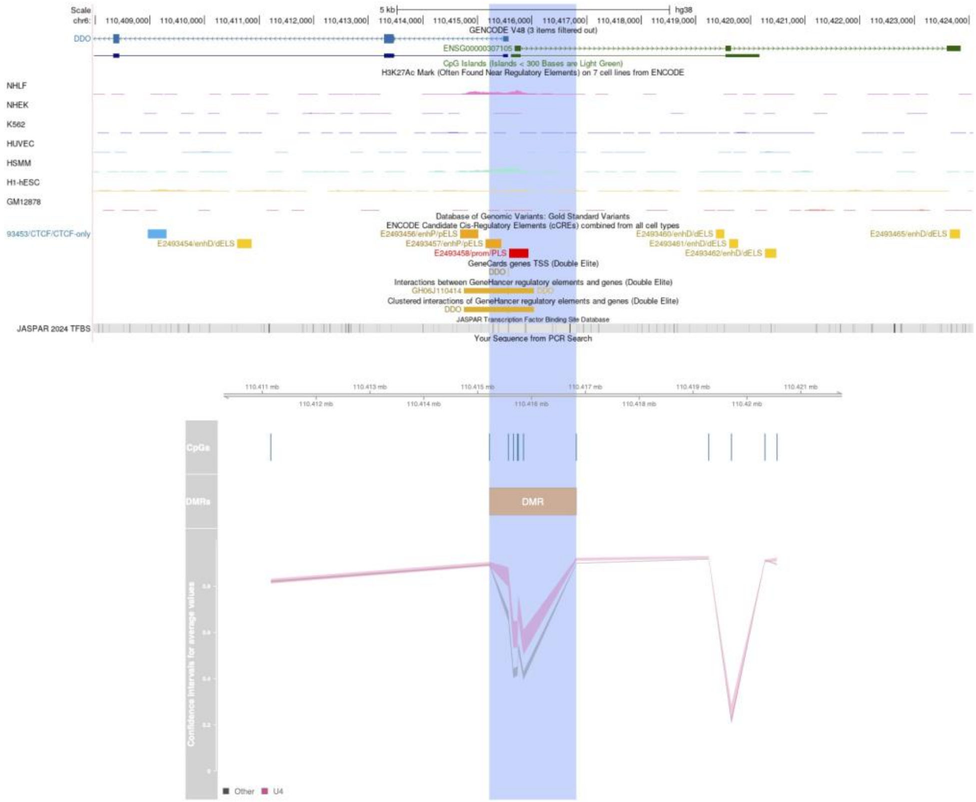

Scale
chr6:
5 kb
hg38
110,409,000 110,410,000 110,411,000 110,412,000 110,413,000 110,414,000 110,415,000 110,416,000 110,417,000 110,418,000 110,419,000 110,420,000 110,421,000 110,422,000 110,423,000 110,424,000
GENCODE V48 (3 items filtered out)
DDO
ENSG00000307105
CpG Islands (Islands < 300 Bases are Light Green)
H3K27Ac Mark (Often Found Near Regulatory Elements) on 7 cell lines from ENCODE
NHLF
NHEK
K562
HUVEC
HSMM
H1-hESC
GM12878
Database of Genomic Variants: Gold Standard Variants
ENCODE Candidate Cis-Regulatory Elements (cCREs) combined from all cell types
93453/CTCF/CTCF-only
E2493454/enhD/dELS
E2493456/enhP/pELS
E2493457/enhP/pELS
E2493458/prom/PLS
E2493460/enhD/dELS
E2493461/enhD/dELS
E2493462/enhD/dELS
E2493465/enhD/dELS
GeneCards genes TSS (Double Elite)
DDO
Interactions between GeneHancer regulatory elements and genes (Double Elite)
GH06J110414
DDO
Clustered interactions of GeneHancer regulatory elements and genes (Double Elite)
DDO
JASPAR Transcription Factor Binding Site Database
JASPAR 2024 TFBS
Your Sequence from PCR Search
110.411 mb 110.413 mb 110.415 mb 110.417 mb 110.419 mb 110.421 mb
110.412 mb 110.414 mb 110.416 mb 110.418 mb 110.42 mb
CpGs
DMRs
DMR
Confidence intervals for average values
Other
U4


*Figure S 12*
